# A *Lifelike* guided journey through the pathophysiology of pulmonary hypertension - from measured metabolites to the mechanism of action of drugs

**DOI:** 10.1101/2023.11.21.23298782

**Authors:** Nathan Weinstein, Jørn Carlsen, Sebastian Schulz, Timothy Stapleton, Hanne Hee Henriksen, Evelyn Travnik, Pär Ingemar Johansson

## Abstract

Pulmonary hypertension (PH) is a pathological condition that affects approximately 1% of the population. The prognosis for many patients is poor, even after treatment. Our knowledge about the pathophysiological mechanisms that cause or are involved in the progression of PH is incomplete. Additionally, the mechanism of action of many drugs used to treat pulmonary hypertension, including sotatercept, requires elucidation. Using our graph-powered knowledge mining software *Lifelike* in combination with a very small patient metabolite data set, we demonstrate how we derive detailed mechanistic hypotheses on the mechanisms of PH pathophysiology and clinical drugs. In PH patients, the concentration of hypoxanthine, 12(S)-HETE, glutamic acid, and sphingosine 1 phosphate is significantly higher, while the concentration of L-arginine and L-histidine is lower than in healthy controls. Using the graph-based data analysis, gene ontology, and semantic association capabilities of *Lifelike*, led us to connect the differentially expressed metabolites with G-protein signaling and SRC. Then, we associated SRC with IL6 signaling. Subsequently, we found associations that connect SRC, and IL6 to Activin and BMP signaling. Lastly, we analyzed the mechanisms of action of several existing and novel pharmacological treatments for PH. *Lifelike* elucidated the interplay between G-protein, interleukin 6, activin, and BMP signaling. Those pathways regulate hallmark pathophysiological processes of PH, including vasoconstriction, endothelial barrier function, cell proliferation, and apoptosis. The results highlight the importance of SRC, ERK1, AKT, and MLC activity in PH. The molecular pathways affected by existing and novel treatments for PH also converge on these molecules. Importantly, sotatercept affects SRC, ERK1, AKT, and MLC simultaneously. The present study shows the power of mining knowledge graphs using *Lifelike*’s diverse set of data analytics functionalities for developing knowledge-driven hypotheses on PH pathophysiological and drug mechanisms and their interactions. We believe that *Lifelike* and our presented approach will be valuable for future mechanistic studies of PH, other diseases, and drugs.

## 1 Introduction

Pulmonary hypertension (PH) is a condition that is defined by a mean pulmonary arterial pressure of more than 20 mmHg at rest [1]. The range of genetic, molecular, and humoral causes that can lead to PH is extensive. PH is classified into five different groups based on clinical and pathological findings as well as therapeutic interventions [2, 3], namely: Pulmonary arterial hypertension (PAH), PH associated with left heart disease (PH-LHD), lung disease (PH-LD), pulmonary arterial obstructions (CTEPH), and PH with unclear and/or multifactorial mechanisms [2, 4, 5]. Pulmonary hypertension (PH) is a condition that affects people of all ages, with a global prevalence of 1%. However, it is more common in individuals over the age of 65. PH is characterized by the remodeling of distal pulmonary arteries, causing a progressive increase in vascular resistance. Vascular remodeling is associated with alterations in vasoconstriction, pulmonary artery-endothelial cell (PAEC), and -smooth muscle cell (PASMC) proliferation, inflammation, apoptosis, angiogenesis, and thrombosis. Some of the lesions found in PH are plexiform lesions, which are characterized by enhanced endothelial cell (EC) proliferation, thrombotic lesions, and the formation of a layer of myofibroblasts, and extracellular matrix between the endothelium and the external elastic lamina [6, 7]. One of the first triggers for the development of PH is thought to be endothelial cell (EC) injury triggering the activation of cellular signaling pathways that are not completely understood [8]. When activated, the endothelium secretes different growth factors and cytokines that affect EC and SMC proliferation, apoptosis, coagulation, attract inflammatory cells, and/or affect vasoactivity to restore homeostasis. Prolonged or chronic activation of the endothelium leads to EC dysfunction, and loss of homeostatic functions [9, 10].

The physiological changes associated with PH are inherently difficult to study in vivo. Limited studies have been carried out on PH patients undergoing lung transplantation, analyzing tissue available in biorepositories, or studying the lungs of deceased patients [11]. Animal models and cultured cells are used extensively in the study of PH. A common approach is to use cultured human cells, including PAECs, PASMCs, and aortic endothelial cells together with animal models. Human ECs are cultivated on plastic with a stiffness five orders of magnitude greater than living tissue [12], exposed to growth media with viscoelastic properties that are different from those of blood [13], and without hemodynamic shear stress [12]. These conditions perturb mechanotransduction and cytoskeletal organization preventing the development of the EC phenotypes found in the human body. Recently, the use of lung-on-a-chip technologies has overcome many of the limitations of human EC cultures [14, 15]. However, even when using these technologies, the results are general and the physiological conditions of a PH patient may differ from those used in a study.

We recently introduced a novel framework for studying the endothelial cell phenotypes of PH patients building on a genome-scale metabolic model (GEM) of the EC which was parameterized by the plasma metabolome from PH patients [16]. We investigated 35 patients with iPAH (n=12), CTD-aPAH (n=12) and CTEPH (n=11) and found significant metabolic differences between the patients. These differences suggest that different disease mechanisms may be involved in each patient. Metabolite concentration measurements constitute a minimally invasive way to understand the physiological state of patients. However, interpreting these results is challenging due to the complex inter-connectivity of molecular networks within cells. Our goal is to identify the relevant biological mechanisms that are active in one patient, or a group of patients based on their metabolic profile. This is very difficult due to the abundance of data and information available in relevant databases and literature. Since Watson, Crick, Franklin, and Wilkins solved the three-dimensional structure of DNA in 1953 [17], advances in genomics [18, 19, 20, 21], RNA sequencing [22], single-cell transcriptomics [23], proteomics [24], metabolomics [25], epigenetics [26], and electronic medical or health records [27] have lead to the creation of a vast amount of biological data. This vast ocean of biological information that grows faster every day, presents many opportunities for the advancement of biology and medicine [28]. However, extracting valuable insights from diverse information sources is very challenging. The obstacles that must be overcome include finding the molecular mechanisms that help us explain the causes for the correlations that we can observe, and enriching the data obtained from a novel research effort with relevant preexisting data. *Lifelike* [29] was developed to help researchers integrate different sources of data, and to explore the molecular mechanisms that help explain experimental observations. In the current manuscript, we use *Lifelike* to elucidate the molecular mechanisms that explain how the differentially expressed metabolites described in [16], are connected to the physiological changes associated with PH.

## 2 Materials and Methods

### 2.1 The data and knowledge mining platform *Lifelike*

*Lifelike* is a software platform for advanced analytics of biological data and knowledge management [29]. *Lifelike* uses a knowledge graph (KG) which consolidates data from several public data sources into a unified single graph database comprising 97,855,162 nodes connected by 162,654,084 edges. The underlying database system is ArangoDB. We have integrated the ontologies Medical Subject Headings (MeSH; https://www.nlm.nih.gov/mesh/meshhome.html) [30] and Gene Ontology (GO) [31, 32], genes and associated taxonomies from NCBI [33, 34], proteins from UniProt [35] and protein-protein interactions from StringDB [36], enzymes from EXPASY [37], chemicals from ChEBI [38], as well as a biomedical data set comprising chemical-gene, chemical-disease, gene-disease and gene-gene relationships derived from text analysis of Medline abstracts [39]. Several conceptually different analyses can be performed on *Lifelike*’s KG as detailed below. A subset of *Lifelike*’s capabilities is available via a public portal https://public.lifelike.bio/ upon self-registration. A guide on how to use *Lifelike* and its data analytics functionalities can be found in the user options panel of the software (bottom left button). *Lifelike* source code is available at https://github.com/SBRG/lifelike.

#### 2.1.1 Annotation of biological entities in text data

*Lifelike*’s rule-based entity recognition system recognizes and highlights eight biological entity types plus user-specified custom annotations. The following entities are detected in text by default: gene, protein, disease, chemical/compound, phenotype/phenomena, anatomy, food, and species. It was utilized to annotate biological entities in text from portable document format files and enrichment tables. The detected entities were used for further downstream analysis, e.g. statistical GO enrichment and semantic analysis.

#### 2.1.2 Enrichment tables

Enrichment tables compile information from *Lifelike*’s knowledge graph into a single table based on a user’s input data set. Using a set of gene names (e.g. from a list of the genes that appear in an article) as an input, *Lifelike* retrieves information from its knowledge graph resources NCBI, UniProt, StringDB, and Gene Ontology, and combines the textual descriptions from each of the databases along with the respective URL in an enrichment table. Enrichment tables were inspected manually or automatically by semantic analysis using *Lifelike*’s word cloud functionality.

#### 2.1.3 Statistical Gene Ontology enrichment

Statistical Gene Ontology (GO) enrichment analysis is used to associate a group of genes with their biological functions. GO enrichment was performed directly from *Lifelike*’s enrichment tables. GO terms were downloaded from https://www.geneontology.org/docs/downloads/ on 10/08/2020. Fisher’s exact test was used to identify GO terms that are statistically enriched in an input gene list. The False Discovery Rate was controlled by the Benjamini-Hochberg procedure [40] and an adjusted p-value (q-value) smaller than the significance level of 0.05 was considered statistically significant.

#### 2.1.4 Semantic analysis

The semantic analysis makes use of *Lifelike*’s text entity recognition system. It can be used on annotated pdf files or enrichment tables. First, enrichment tables were generated using a list of genes as input (e.g. genes obtained from initial radiate analysis). Then, the textual descriptions from *Lifelike*’s KG are analyzed to detect and extract biological entities, which include genes, that are semantically associated with the input (See section 2.1.1). The extracted genes were used for several downstream analyses, either for statistical GO enrichment or as an input for another iteration of enrichment table generation and semantic analysis.

#### 2.1.5 Semantic entity clouds

Entity clouds are a facile way to grasp higher concepts from text quickly. They were generated in *Lifelike* from pdf files and enrichment tables. Biological entities (e.g. proteins, compounds, etc) were automatically detected in text, counted (either in the entire table or row-wise), and visually displayed as word clouds. The higher the count of an entity, the larger its size is in the entity cloud. These are especially useful for identifying associated chemicals, phenotypes, proteins, and genes, which can be used to create enrichment tables.

#### 2.1.6 Generation of knowledge maps

Knowledge maps provide a visual storyboard of results, incorporating elements such as images, links to related maps, PDFs, enrichment tables, graphical files, office documents, and biological entities. It is also possible to drag annotated entities from enrichment tables or annotated pdf files onto a map and search across the literature. Results from data analyses were summarized on knowledge maps to capture, interpret, and contextualize data analysis results and the knowledge gained. The elements on a knowledge map were linked to their origins, i.e. to URLs of enrichment tables, pdf, and other files in *Lifelike* were saved as attributes of a knowledge map element.

### 2.2 Graph Data Science

We conducted knowledge mining utilizing graph algorithms on a modified version of the curated graph database Reactome, which is a manually curated, peer-reviewed pathway database that is open-source, and open-access [41, 42]. The graph data science (GDS) code contains libraries that allow us to find molecules that affect and are affected by genes and then analyze their connections in a graph database. Source code for graph data science is available at https://github.com/SBRG/GDS-Public/tree/main. Different graph algorithms were run (detailed below in sections 2.2.3 and 2.2.4), including radiate analysis (Radiate analysis notebook) with subsequent generation of Sankey visualizations (Radiate traces notebook) and all shortest paths (Shortest path notebook).

#### 2.2.1 Construction of the Reactome graph database

The Reactome database [41, 42] was downloaded as a neo4j graph database (https://reactome.org/download-data version 75). A series of database queries was used to generate a database version suitable for graph data science which can be followed in detail in the provided Jupyter notebook (Reactome processing notebook). Briefly, nodes, labels, and relationships not required for graph algorithmic analyses were removed. For instance, this included nodes like person, affiliation, and taxa as well as all nodes representing entities of organisms other than *Homo sapiens*. Sub-cellular locations (compartments) of biological entities were set as node properties. To allow for improved graph traversal, selected relationships were reversed or added. Because currency metabolites, e.g. ATP, NAD(P)H and H^+^, can artificially connect metabolic reactions and pathways in network analyses [43, 44] we labeled such compounds plus the regulatory protein ubiquitin accordingly (Currency nodes) and thereby excluded them from all analyses. Finally, the database was transformed into an ArangoDB graph database consisting of 1,703,054 nodes and 3,368,926 edges, available at [29]. Furthermore, we allowed only 12 selected edge types in the database to be traversed by the graph algorithms (Supplementary Table 1).

**Table 1:** Radiate analysis, enrichment tables, traces, and knowledge maps. Each entry in the resource column is a link.

| Number | Resource |
| --- | --- |
| 1 | Initial radiate analysis |
| 2 | The metabolites influence $G\alpha(i)$ |
| 3 | Genes related to SRC |
| 4 | Genes related to SRC expanded |
| 5 | S1P, L-Glu, Hyp, and 12(S)-HETE augment IL6 expression |
| 6 | SRC upregulates IL6 expression |
| 7 | Genes related to IL6 |
| 8 | Shortest path from IL6 to STAT3 |
| 9 | Radiate analysis using the complex formed by SMAD2, SMAD3, and SMAD4 as sources |
| 10 | Genes influenced by SMAD2 and SMAD3 |
| 11 | Shortest paths from a SMAD3 complex to cell cycle regulators |
| 12 | Radiate analysis using Activin A and Activin B as sources |
| 13 | Protein targets of Activin A and Activin B |
| 14 | Radiate analysis using the complex formed by SMAD1, SMAD5, or SMAD9, and SMAD4 as sources |
| 15 | Genes that influence SMAD1, SMAD5, or SMAD9, and SMAD4 |
| 16 | Radiate analysis using BMP2 as the source |
| 17 | Gene targets of BMP2 |
| 18 | BMP2 semantic iterative analysis results |
| 19 | BMP9 semantic iterative analysis results |
| 20 | Journey storyboard |
| 21 | Analyzing and enriching articles and figures |
| 22 | Pathways in detail |

#### 2.2.2 Finding the correct nodes for GDS analysis

Graph algorithms require finding specific molecules in the graph. Source and/ or target node sets (e.g. 6 differential metabolites from pulmonary hypertension patients) were found in the graph using Reactome standard identifiers (stId; obtained from the Reactome website: https://reactome.org). The Reactome database models metabolites and other node types depending on their subcellular location (multiple cellular compartments are possible). Unless stated otherwise, metabolite nodes where found in the Reactome graph by selecting the extracellular compartment which allows the investigation of the effect of extracellular plasma metabolites on cellular entities like receptors and downstream effectors.

#### 2.2.3 All shortest paths

To calculate all possible shortest paths between a source and a target node set, we developed a custom Python script that uses NetworkX’s all_shortest_paths method (https://networkx.org/). Shortest path calculations were performed either on an unweighted or a weighted network (for weighting details see section 2.2.4). In unweighted networks, a shortest path between a source and a target node is defined as the path with the minimum number of edges connecting them. In weighted networks, a shortest path between a source and a target node is defined as the path with the minimum sum of edge weights along a path connecting them. This analysis yielded files that were manually uploaded to *Lifelike* for interactive visualization and detailed analysis.

#### 2.2.4 Radiate analysis and tracing

To measure the importance of network nodes with respect to an input data set, we used a two-step approach which we termed “Radiate analysis and tracing”. This approach is an exploratory graph analysis method where only one input data set is used. First, ranking of network nodes with respect to an input data set is achieved using personalized Pagerank (PPR) [45, 46]. PPR has been used previously for the analysis of biological networks including protein-protein interaction and metabolic [47] as well as gene-regulatory networks [48, 49]. An input data set used as sources (e.g. differential metabolites) was mapped on the graph and PPRs of network nodes were calculated considering the edge directionality of the network. Forward radiate analysis uses the edge directionality in the network as is and answers the question “Which nodes in the network are highly influenced by the source input data set?”. In reverse radiate analysis, the directions of all edges in the graph were temporarily reversed prior to calculation of PPRs. It answers the question “Which nodes in the network have the highest influence over the source input data set?”. We used the NetworkX implementation of Pagerank (NetworkX documentation) which allows for personalization of the algorithm using an user-defined source data set. The obtained probability distribution was normalized such that the sum of all pageranks in the network (including the source nodes) equals 1. This step returned a ranked list of network nodes for each forward and reverse radiate analysis where the Pagerank of a node measures its importance with respect to the input data set. Second, trace graphs from source nodes to selected target nodes identified by radiate analysis were generated. This allowed us to visualize and to understand the detailed connectivity of source nodes used as algorithmic input to target nodes identified by radiate analysis. Trace graphs were generated using either all shortest paths on the unweighted network (see above) or paths of highest influence on a weighted network. Weighting of each edge in the network was done using the inverse of the maximum PPR from one of the two nodes connected by the edge. Paths of highest influence were found by calculating all shortest paths on the weighted network. The shortest paths contain the smallest number of edges while the paths of highest influence have the minimum sum of inverse PPR values. This step yielded a Sankey file which is uploaded to *Lifelike* for detailed analysis and interpretation.

### 2.3 Experimental design

The individual data analytics functionalities of *Lifelike* can be combined in various ways to build mechanistic knowledge from input data sets as shown in Figure 1. In the present study, we followed an analysis workflow as depicted in Supplementary Figure 1 with the aim of generating deep knowledge from a comparatively small set of input data. Our analysis workflow started with a radiate analysis using the 6 differential metabolites as input. We performed both forward and reverse radiate analysis, to identify network nodes that are highly relevant with respect to the metabolite input data set. We mainly focused on highly relevant protein nodes identified using forward radiate analysis, unless stated otherwise. To understand and interpret the connectivity of metabolites to identified network nodes (e.g. proteins), we generated interactive Sankey diagrams, where each path can be inspected in a very detailed manner by the user. *Lifelike* enrichment tables were generated using lists of highly affected genes. Enrichment tables allow us to perform two downstream analysis tasks: semantic and statical enrichment analysis. Semantic analysis yielded word clouds highlighting different biological entity types which are highly mentioned in the associated enrichment tables. Statistical enrichment analysis against gene ontology terms identified biological processes affected by the 6 plasma metabolites. To further expand the generated knowledge, semantic analysis was performed iteratively. All analyses results, including interpretations and knowledge gained were summarized on *Lifelike* knowledge maps which represent an effective tool for data storytelling.

**Figure 1:**
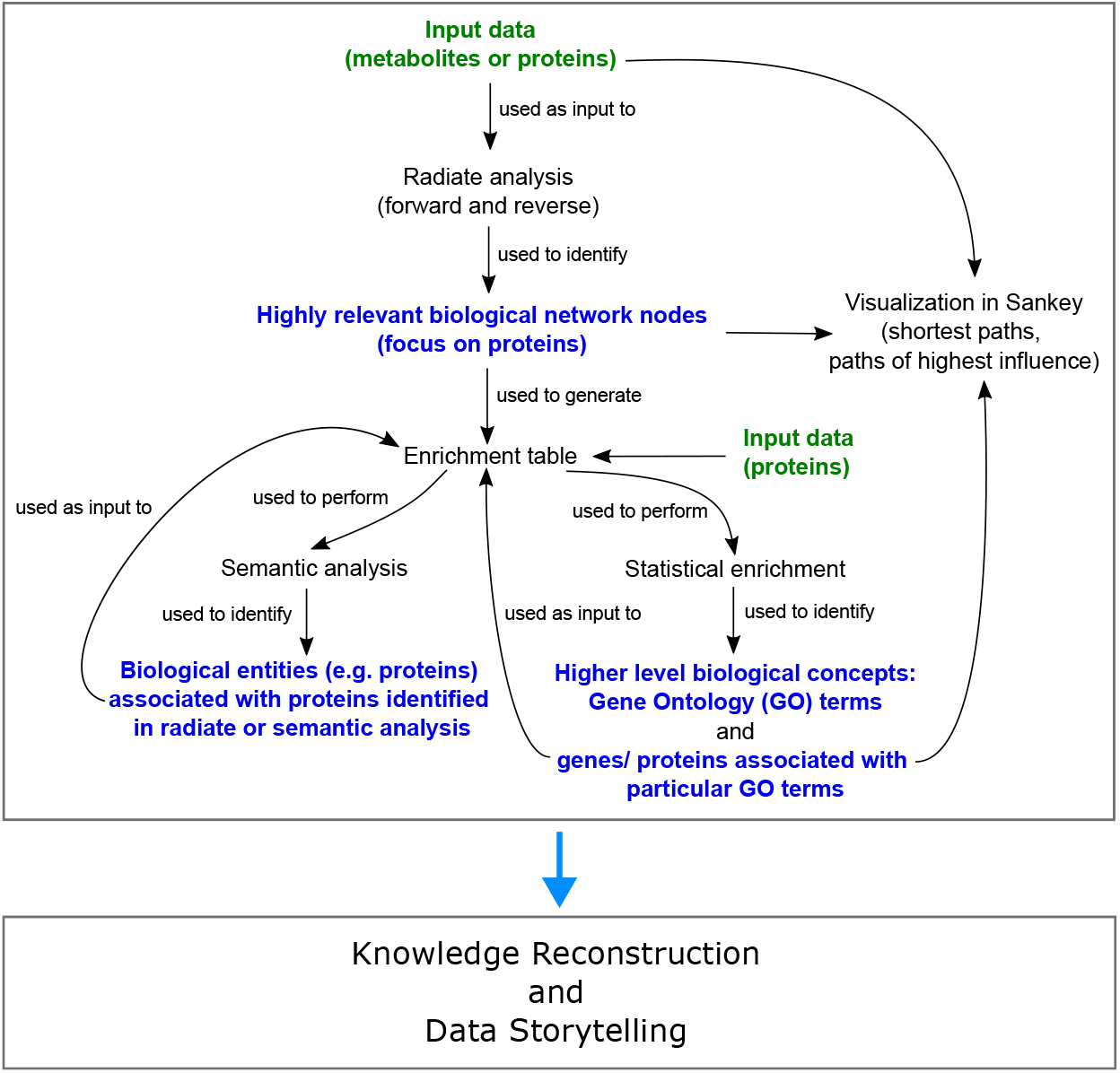
Schematic workflow employing *Lifelike*’s functionalities for the generation of deep knowledge from as little as 6 metabolites detected in the plasma of pulmonary hypertension patients.

## 3 Results

### 3.1 S1P, 12(S)-HETE, and L-Glu are more abundant in PH patients contributing to increased endothelial barrier permeability, vasoconstriction, and vascular cell proliferation

We found that the average concentrations of hypoxanthine (Hyp), S 12-Hydroxyeicosatetraenoic acid (12(S)-HETE), L-glutamic acid (L-Glu), and sphingosine 1 phosphate (S1P) increased more than two-fold in treated PH patients compared to the control group. Furthermore, the measured concentration of L-Arginine (L-Arg) and (L-Histidine) L-His are significantly lower in treated patients than in the control group (less than 71%) [16]. We therefore searched for the molecules that are influenced by the metabolites as described in section 2.2.4 using radiate analysis. The activation of Gi protein complexes by a G-protein-coupled receptor (GPCR) upon binding its corresponding ligand and the non-receptor tyrosine kinase SRC, were identified as crucial targets for S1P, 12(S)-HETE, and L-Glu (Table 1 rows 1,2). GPCRs are important mediators of cellular signaling that are involved in the regulation of vascular cell proliferation [50], vascular tone [51, 52, 53, 54, 55, 56] and endothelial barrier function [57, 58]. Diverse G protein complexes, such as Gi/0, Gq, G12, and G13, enable GPCRs to elicit cell and tissue-specific responses to ligands.

#### 3.1.1 Normally S1P preserves endothelial barrier function (Figure 2a)

In healthy individuals, the normal plasma concentration of S1P (0.1 to 0.8 μM) is sufficient to activate the GPCR S1PR1 in PAECs [59]. S1PR1 signals through Gi/0, PI3K and AKT1 to preserve endothelial barrier function. AKT1 causes cytoskeletal remodeling and EC spreading through RAC1 and Cortactin. Additionally, AKT1 activates eNOS, which catalyzes the production of NO, which causes the concentration of cGMP to increase. Then cGMP activates PKG I, which prevents the release of Ca2+ from the endoplasmic reticulum and activates MLCP. Subsequently MLCP deactivates MLC preventing cell contraction [51, 54, 60, 61, 62](Figure 2a, and Supplemental section 1.1.1).

**Figure 2:**
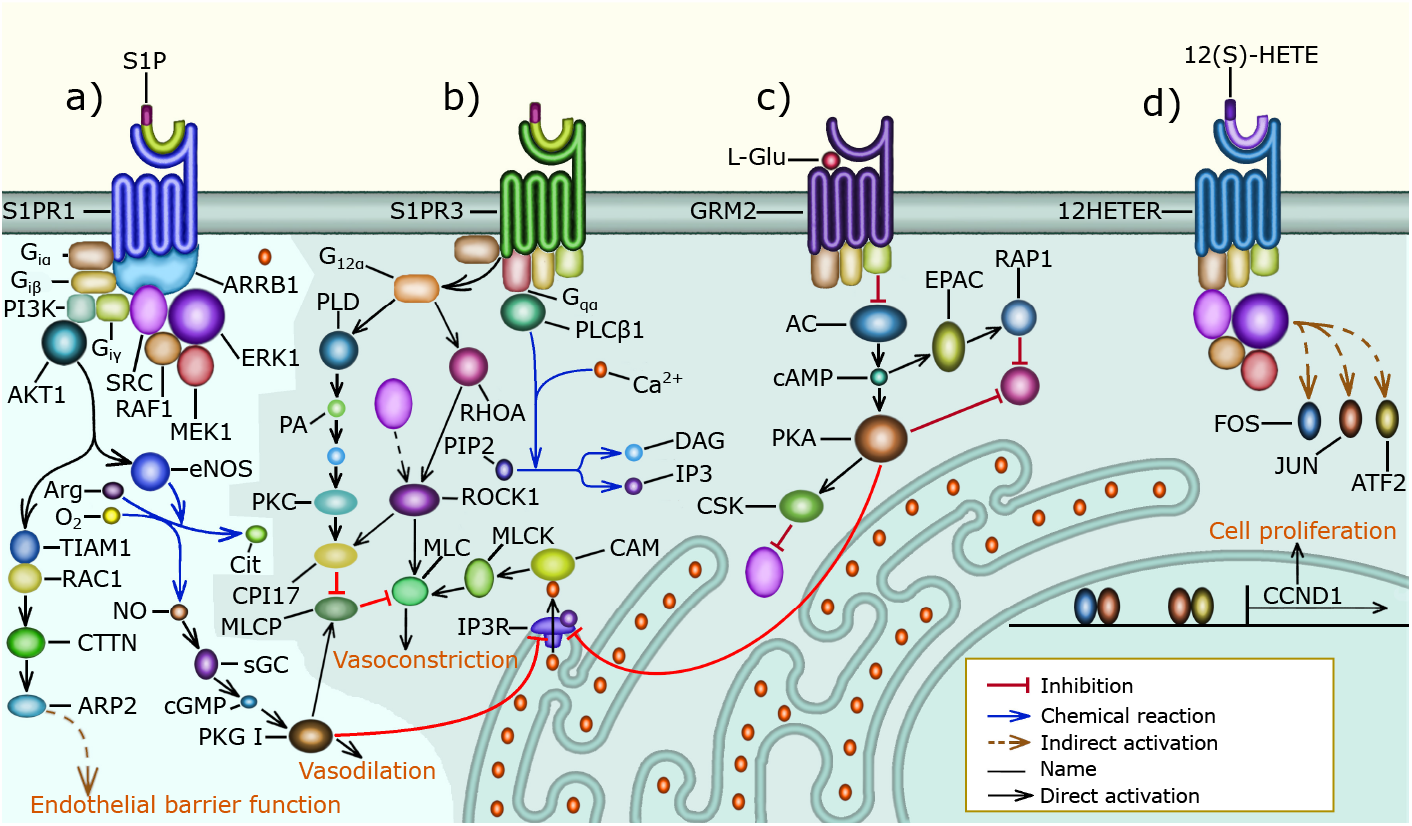
G protein signaling in pulmonary vascular cells: The shaded area of the figure includes signaling pathways that are overactivated in PH patients, a) Normally, S1P binds S1PR1 and preserves endothelial barrier function through the activation of the Gi, PI3K, AKT, and RAC1 signaling pathway that causes PAECs to increase the surface they cover through cytoskeletal remodeling. Gi, PI3K, and AKT signaling also activate eNOS to increase NO production and secretion. NO activates sGC increasing cGMP availability leading to PKG I activation. PKG I prevents MLC activation and cell contraction by decreasing the Ca2+ concentration in the cytosol and by activating MLCP. Lastly, S1P promotes normal PAEC proliferation through Gi, ARRB1, SRC, and ERK signaling, which promotes Cyclin D1 expression, b) In PH patients, the concentration of S1P in the plasma is elevated and this activates SIPR2 (Not shown), and S1PR3 signaling resulting in MLC activation and cell contraction. S1P3 increases MLC activity through the G12, PKC, and MLCP signaling pathway, the G12, RHOA, and ROCK signaling pathway that requires SRC activity, and the Gq, PLC*β*, Ca, and MLCK signaling pathway. In PAECs, MLC activation results in cell rounding and endothelial barrier permeability. Additionally, in PASMCs MLC activation leads to vasoconstriction, c) The elevated concentration of L-Glu in the plasma of PH patients activates GRM3 (Not shown) and GRM2-mediated Gi signaling that inhibits AC causing the cAMP concentration to decrease. The absence of cAMP inhibits PKA, allowing the release of Ca2+ from the sarcoplasmic reticulum. Also, the decrease in cAMP leads to an increase in SRC activity. Further, the reduction in cAMP increases RHOA activity, which causes MLC activation, loss of endothelial barrier function, and vasoconstriction, d) The concentration of 12(S)-HETE is abnormally high in many PH patients, 12(S)-HETE binds 12HETER, leading to the activation of the Gi, ARRB1, SRC, and ERK signaling pathway, which causes excessive vascular cell proliferation. Additionally, 12HETER can signal through Gq and G12 causing PAEC contraction (not shown).

#### 3.1.2 In PH patients S1PR2, S1PR3, 12HETER, GRM2, and GRM3 act in concert to promote vascular cell proliferation, inhibit apoptosis, and increase the activity of MLC resulting in vasoconstriction (Figure 2b-2d)

The S1P receptors S1PR2, and S1PR3 [51, 54, 59, 61], and the 12(S)-HETE receptor 12HETER [50, 58, 63] are present in the cell membrane of PAECs and PASMCs. S1PR2, and S1PR3 are only activated when the concentration of S1P is abnormally elevated as observed in most PH patients. Similarly, the activity of 12HETER is abnormally elevated in PH patients. S1PR2, S1PR3 (Supplementary section 1.1.2, and Figure 2b) and 12HETER (Supplementary section 1.1.4, and Figure 2d) signal through Gq, PLC*β*, IP3, and DAG. IP3 promotes the release of Ca2+ from the endoplasmic reticulum, and DAG inhibits MLCP, resulting in MLC activation [61, 64, 65]. Additionally, S1PR2, S1PR3 and 12HETER also activate G12, and G13 signaling, which causes RHOA -mediated MLC activation [52, 54, 55] and preserves MLC activity through the inhibition of MLCP [53]. Moreover, the elevated concentration of L-Glu found in PH patients suffices to activate GRM2, and GRM3, which are expressed by PAECs [66, 67]. GRM2, and GRM3 signal through G i/0 to decrease the concentration of cAMP, which inhibits the release of Ca2+ from the endoplasmic reticulum and is required for the RAP1-dependent inhibition of RHOA. Therefore, GRM2, and GRM3 augment MLC activity by decreasing the concentration of cAMP [68, 69, 70] (Figure 2c, Supplemental section 1.1.3). In PAECs MLC activity increases endothelial barrier permeability. Furthermore, in PASMCs MLC activity leads to vasoconstriction. Additionally, S1PR2, S1PR3, 12HETER, GRM2, and GRM3 signal through Gi/0 and SRC resulting in the proliferation of PAECs and PASMCs (Figure 2d and Supplemental section 1.1).

#### 3.1.3 SRC is an important mediator of S1PR1, S1PR3, GRM2, GRM3, and 12HETER signaling in PH patients that stimulates excessive vascular cell proliferation, increases endothelial barrier permeability, and causes vasoconstriction (Figure 2b-2d)

The non-receptor tyrosine kinase SRC was the first proto-oncogene to be described and is involved in the regulation of cell growth, differentiation, and survival [71]. SRC plays an important role in the pathophysiology of PH and is a potential drug target for the treatment of PH [72]. SRC is an important mediator of S1PR1, S1PR3, GRM2, GRM3, and 12HETER signaling because in PH patients, overactive Gi/0 signaling leads to excessive PAEC and PASMC proliferation by activating the SRC/ERK signaling pathway that induces the expression of Cyclin D1 [50, 73, 74, 75] (Figure 2d, Supplemental section 1.1.4). Additionally, in a rat model SRC mediates hypoxic pulmonary vasoconstriction through the translocation of ROCK, which is necessary for MYPT-1 activation. MYPT-1 inhibits MLCP resulting in increased MLC phosphorylation and vasoconstriction [76]. Furthermore, SRC phosphorylates VE-cadherin to regulate endothelial barrier permeability in response to flow-mediated mechanosensory stimuli, and in PH patients this leads to internalization of VE-cadherin and increased barrier permeability [77]. The molecular mechanisms mentioned above imply that SRC is an essential component of many pathways that connect the differentially expressed metabolites with vasoconstriction, endothelial dysfunction, and vascular cell proliferation. We call molecules like SRC convergence nodes.

### 3.2 IL6 signaling promotes pulmonary vascular cell proliferation and increases endothelial barrier permeability in PH patients

#### 3.2.1 SRC activates the expression of IL6, leading to autocrine and paracrine IL6 signaling (Figure 3j)

Due to the importance of SRC as a convergence point for GPCR signaling, we performed a semantic analysis on textual descriptions derived from public databases such as NCBI, Uniprot, and StringDB, resulting in an enrichment table with related genes (Table 1 row 3), which we further analyzed utilizing the Gene Ontology (GO). This identified IL6 signaling, which is associated with STAT1, IL6, CBL, STAT3, and SRC. After that, we traced the shortest paths from SRC to IL6, which involve the AKT signaling pathway, which induces IL6 expression through CREB1 activation, and the RAS signaling pathway, which induces IL6 transcription through CEBPB phosphorylation (Table 1 rows 5,6). This is in accordance with experimental observations that SRC activates the expression of IL6 [78, 79]. The diverse mechanisms that allow SRC to increase IL6 expression likely contribute to the elevated concentration of IL6 found in the serum and lungs of PH patients [80, 81] (Figure 3j). In alignment with this, overexpression of IL6 triggers PH and right ventricle hypertrophy in mice by promoting pulmonary arterial muscularization and the formation of occlusive lesions composed of endothelial cells and T-lymphocytes in the distal pulmonary arterioles [82]. Also, higher concentrations of IL6 are associated with a greater risk of death [80].

**Figure 3:**
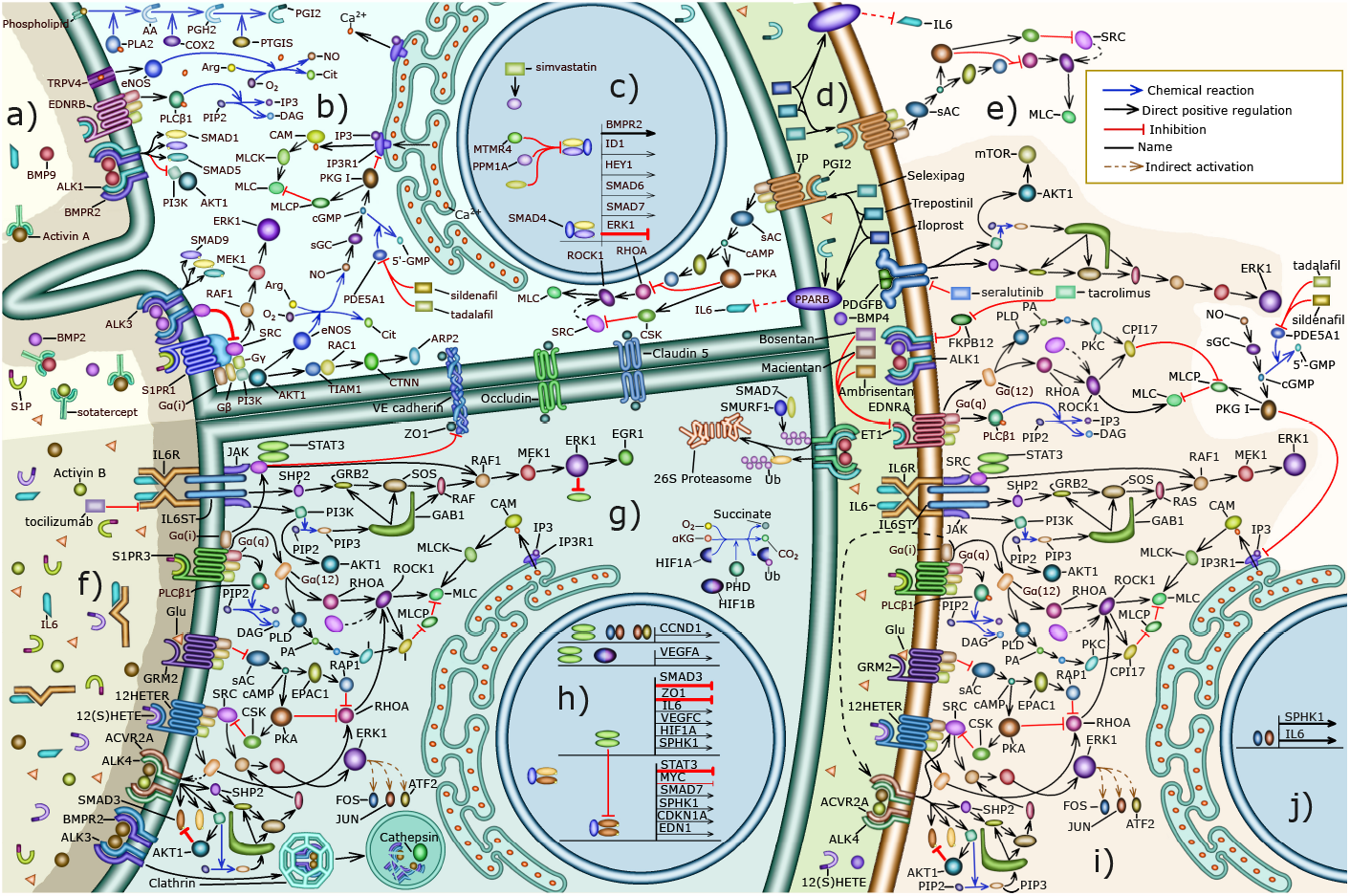
Summary of the signaling pathways involved in the pathophysiology of PH and the mechanisms of action of drugs that we explored: a) The lumen of a pulmonary blood vessel of a treated PH patient, b) The cytosol of a PAEC in a healthy person or treated PH patient, c) The nucleus of a PAEC in a healthy person or treated PH patient, d)The basement membrane separating PAECs from PASMCs, e) Mechanisms that cause vasodilation in healthy PASMCs, f) The lumen of a pulmonary blood vessel from a PH patient, g)The cytoplasm of a PAEC from a PH patient, h) The nucleus of a PAEC from a PH patient, j) The mechanisms that cause PASMC contraction and proliferation, which are overactive in PH patients, j) The nucleus of a PASMC from a PH patient.

#### 3.2.2 IL6 signaling increases endothelial barrier permeability, inhibits apoptosis, and increases cell proliferation through the activation of STAT, ERK and AKT signaling (Figure 4a-4f)

. The importance of IL6 in PH led us to perform a semantic analysis of STAT1, IL6, CBL, STAT3 and SRC that resulted in an enrichment table that contains genes related to IL6 (Table 1 row 7). Subsequently, we used GO enrichment to associate the genes related to IL6 with biological mechanisms that are important for the pathophysiology of PH, including positive regulation of ERK1 and ERK2 signaling, positive regulation of smooth muscle and endothelial cell proliferation, negative regulation of apoptotic process, and platelet activation. The IL6 receptor, IL6R is not expressed in PAECs. However, in the plasma of PH patients IL6 binds the soluble form of its receptor (sIL6R), and then both bind the coreceptor IL6ST (gp130) present in the cell membrane of PAECs forming a hetero-hexamer complex that initiates trans IL6 signaling [80, 81]. IL6 signaling leads to the activation of STAT signaling, ERK signaling, and AKT signaling (Figure 4a-4c) [83]. IL6 signaling, through ERK1, ERK2, and STAT3 promotes PAEC and PASMC proliferation by augmenting the expression of Cyclin D1, MYC, and PIM1 (Figure 4a, and Supplementary section 1.2.2) [81, 84]. Moreover, SRC and STAT3 are activated by IL6 signaling and increase endothelial barrier permeability by causing the internalization of adherence and tight junction components including VE-cadherin and ZO1 [85, 86, 87] (Figure 4d, and Supplementary section 1.2.3). Additionally, IL6 signaling integrates inflammatory and hypoxic signals to increase VEGFA [88] expression, which further increases endothelial barrier permeability (Figure 4f, and Supplementary section 1.2.4). Further, IL6-activated AKT signaling promotes PAEC survival by stimulating the expression of Survivin [89] and suppressing BAX-mediated apoptosis [90] (Figure 4c, and Supplementary section 1.2.5).

**Figure 4:**
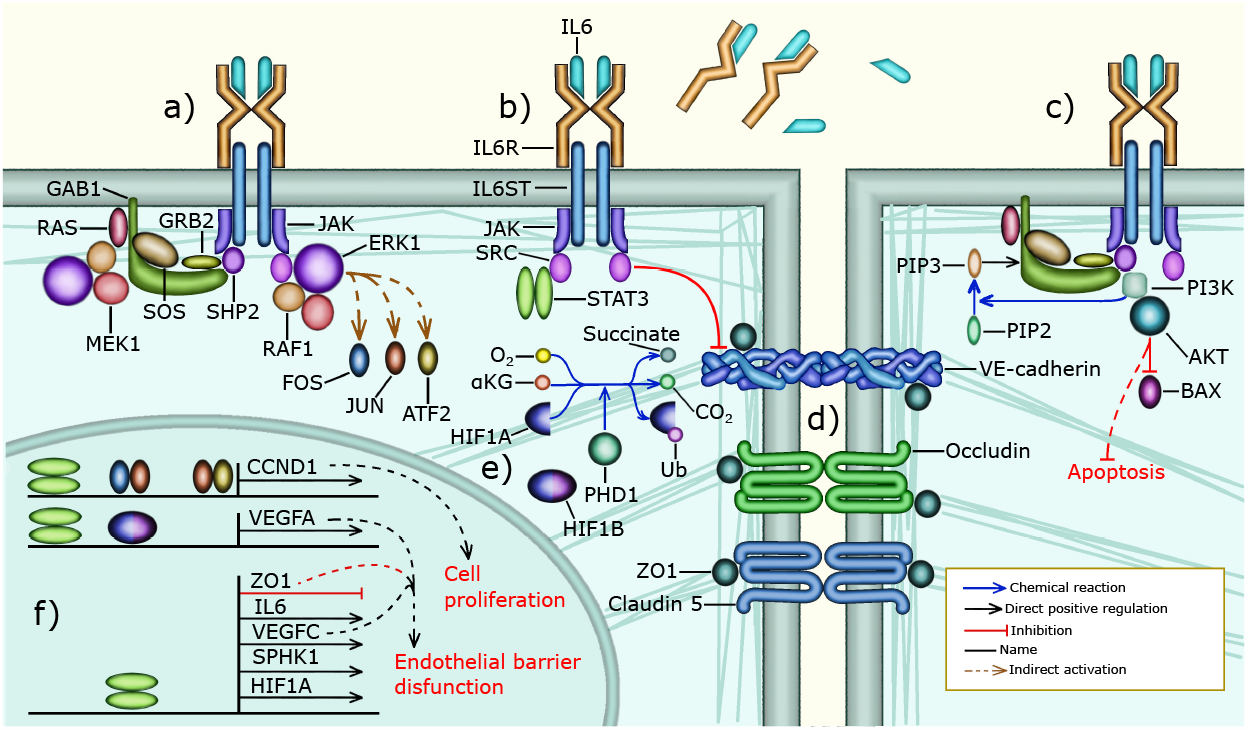
IL6 signaling in the endothelium of a PH patient: a) PAECs do not express IL6R. However, sIL6R and IL6 are present in high concentrations in the blood plasma of PH patients. The complex formed by IL6, sIL6R and IL6ST activates JAK and then SHP2 or SRC to activate ERK signaling. SRC directly activates RAF1, leading to the activation of MEK1, and ERK1. SHP2 on the other hand requires GRB2, SOS, and RAS to activate RAF1, MEK1, and ERK1 signaling, b) IL6 signaling can also activate STAT3 through SRC. Additionally, SRC phosphorylates VE-cadherin, c) IL6 signaling can also activate PI3K through SHP2 and GAB1. PI3K then phosphorylates PIP2 to produce PIP3 which is needed to recruit GAB1 near the cell membrane. Additionally, PI3K activates AKT1, which inhibits BAX-mediated apoptosis, d) STAT3 forms a dimer that inhibits the transcription of ZO1 resulting in the internalization of adherence and tight junctions increasing endothelial barrier permeability, e) Hypoxia prevents the ubiquitination and degradation of HIF1A. Then HIF1A binds HIF1B forming a complex that regulates the transcription of several genes, f) STAT3, FOS, JUN, and ATF bind the promoter of CCND1 to activate the expression of Cyclin D1 allowing G1/S cell cycle progression, which leads to PAEC proliferation. IL6 is involved in some of the mechanisms through which hypoxia triggers PH, for instance HIF1A, HIF1B and STAT3 bind to the VEGFA promoter and activate the expression of VEGFA. Additionally, STAT3 activates the expression of IL6, VEGFC, SPHK1, and HIF1A. STAT3 also inhibits the transcription of ZO1.

### 3.3 In PH patients, activin signaling and cell proliferation are overactivated while BMP signaling and apoptosis are inhibited

In our SRC-centered iterative semantic analysis we encountered GDF2(BMP9), SMAD1, SMAD3, and SMAD4 (Table 1 4). Additionally, SMAD2 and SMAD3 are intermediary nodes in the shortest path trace from IL6 to STAT3 (Table 1 8). The TGF*β* family of signaling pathways that include activin signaling and BMP signaling is involved in the regulation of EC identity, motility, and barrier function [91, 92]. Further, BMPR2, ALK1, ENG, and SMAD9 mutations are associated with PH [93] and the activin and BMP signaling pathways are involved in the pathophysiology of PH [94, 95]. Therefore, we decided to further explore the effect of activin and BMP signaling in PH.

#### 3.3.1 In PH patients, SMAD2 and SMAD3-mediated activin signaling increases the production of S1P and ET1, inducing vascular cell proliferation and vasoconstriction (Figure 5g). Additionally, SMAD-independent activin signaling stimulates ERK signaling (Figure 5b) and causes BMPR2 internalization and degradation, further increasing vascular cell proliferation (Figure 5h)

Activin A is overexpressed in the PAECs of PH patients [96]. The canonical effect of activin signaling is the phosphorylation and activation of SMAD2 and SMAD3 [96, 97, 98]. Therefore, we carried out a radiate analysis using the complex formed by SMAD2, SMAD3, and SMAD4 as the source (Table 1 row 9). Then, we took the genes targeted by SMAD2, SMAD3, and SMAD4 signaling, created an enrichment table (Table 1 row 10), searched for the GO biological processes that are significantly associated with the genes and found several GO terms associated with cell cycle regulation.

**Figure 5:**
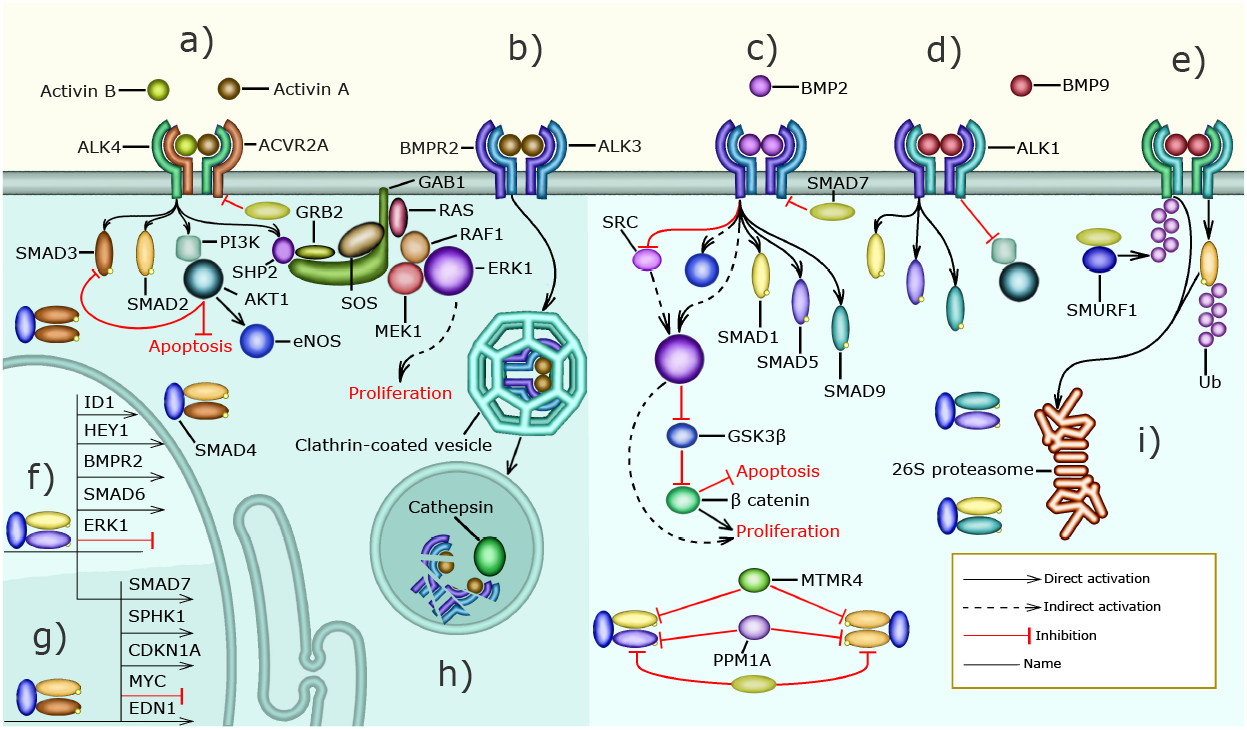
Activin and BMP signaling: The shaded areas highlight pathways that are overactivated in PH patients a) The concentration of activin A and activin B in the serum of PH patients is higher than normal. Activin A and activin B bind ACVR2A with a higher affinity than BMP2 and other TGF*β*-family ligands. When an activin dimer (AA, BB, or AB) forms a complex with ACVR2A and ALK4, it can activate SMAD2, SMAD3, AKT, and ERK signaling. AKT inhibits apoptosis and ERK allows vascular cell division leading to excessive vascular cell proliferation, b) Activin A also binds BMPR2 with high affinity, preventing BMP2 signaling through competitive inhibition, c) In healthy subjects, BMP2 binds BMPR2 and ALK3 resulting in the activation of SMAD1, SMAD5 and SMAD9. Additionally, when BMP2 binds BPMR2 it inhibits SRC. Further, BMP2 signaling regulates apoptosis and promotes normal endothelial proliferation through the ERK-mediated activation of *β* catenin d), BMP9 can form a complex with BMPR2 and ALK1 resulting in the activation of SMAD1, SMAD5, and SMAD9. Additionally, BMP9 inhibits PI3K and AKT signaling through BMPR2 e) Further, BMP9 can bind aCVR2A and ALK1 forming a complex that leads to the activation of SMAD2. SMAD7 competes with SMAD2 for the receptor. Moreover, SMAD7 recruits and collaborates with the E3 ligase SMURF1 that ubiquitinates active R-SMADS and TBRIs targeting them for 26S proteosome-mediated degradation, BMP-activated SMAD1, SMAD5, and SMAD9 can form complexes with SMAD4 that are transported into the nucleus where they inhibit the expression of ERK1 and activate the expression of HEY1, SMAD6, SMAD7, and ID1, Activin-induced SMAD2 and SMAD3 form complexes with SMAD4 that are transported to the nucleus where they inhibit the expression of MYC and activate the transcription of SMAD7, SPHK1, and CDKN1A, h) Actvin A binding leads to the internalization of BMPR2 and ALK3 in Clathrin-coated vesicles that fuse with lysosomes present in the cytoplasm. The lysosomes contain Cathepsin peptidases that are active due to the low pH and degrade BMPR2 and ALK3, i) SMAD6, SMAD7, PPM1A, and MTMR4 downregulate the formation of SMAD transcription factor complexes.

After that, we traced the shortest paths that connect the SMAD complex to important cell cycle regulators (Table 1 row 11). We found that SMAD3 signaling activates the expression of CDKN1A (p21) and downregulates MYC directly (Figure 5g) and cyclin D1 indirectly, therefore preventing cell proliferation. This is in accordance with what has been observed experimentally in many cell types including human umbilical vein ECs [99, 100] (Supplementary section 1.3.1.1 and Figure 5g).

In PASMCs, activin signaling stimulates excessive proliferation [98] and this may be explained by SMAD-independent pathways stimulated by activin signaling. Therefore, we performed a radiate analysis using activin A and B as the sources (Table 1 row 12). The GO terms significantly associated with the genes targeted by activin A and B (Table 1 row 13) include negative regulation of apoptotic process, positive regulation of cell population proliferation, and negative regulation of BMP signaling pathway supporting the assumption above.

Additionally, we used the entity cloud functionality of *Lifelike* on the enrichment table above to highlight frequently mentioned genes associated with activin signaling, including AKT1, BAD, BAX, SRC, ERK, RHO, and RAC1, which are involved in SMAD-independent activin signaling (Supplementary section 1.3.1.2).

It has been reported in the literature that activin upregulates AKT [101] and ERK [102, 103, 104] signaling and induces the internalization and degradation of BMPR2 (Supplementary section 1.3.1.2, Figure 5a, 5b, 5h). AKT inhibits apoptosis and prevents the phosphorylation and nuclear translocation of SMAD3 [105]. Further, ERK signaling causes excessive proliferation in the PAECs of PH patients [106, 107, 108].

Literature curation identified two possible mechanisms through which SMAD-dependent activin signaling is detrimental for PH patients. In PASMCs SMAD2, SMAD3, and SMAD4 stimulate the expression of SPHK1, an enzyme that catalyzes S1P production [109] contributing to the excessive PASMC proliferation and vasoconstriction experienced by PH patients and it could be speculated that this also occurs in PAECs leading to an increase in endothelial barrier permeability. Furthermore, activin A augments the production of endothelin-1 (ET1) leading to vasoconstriction [98]. The promoter of EDN1, which encodes a preproprotein that is proteolytically processed to generate ET1, contains a SMAD binding site, and phosphorylated SMAD3 activates the transcription of EDN1 in cultivated HUVECs [110].

#### 3.3.2 Under normal conditions, BMP2 and BMP9 activate SMAD1, SMAD5 and SMAD9, which promote the transcription of BMPR2 and inhibit the transcription of ERK1 and ERK2. Additionally, BMP2-activated BMPR2 inhibits PAEC proliferation by inhibiting SRC activation (Figure 5c-5f)

BMP signaling canonically leads to the phosphorylation of SMAD1, SMAD5, and SMAD9 [92, 111]. We performed a radiate analysis with these R-SMADS as sources (Table 1 row 14). Then, we created an enrichment table with the most significant proteins that target canonical BMP signaling (very few proteins are targeted by these SMADs) (Table 1 row 15). After that, using GO we found that: negative regulation of cell population proliferation, regulation of smooth muscle cell proliferation, BMP signaling pathway, and activin receptor signaling pathway are significantly associated with the genes that regulate SMAD1, SMAD5, and SMAD9.

Similarly, to study the effect of both SMAD-dependent and SMAD-independent BMP signaling we performed a radiate analysis using BMP2 as the source (Table 1 row 16). Then we created an enrichment table containing the gene targets of BMP2 (Table 1 row 17). After that, using GO we found that the targets of BMP2 are significantly associated with the following terms: activation of cysteine-type endopeptidase activity involved in apoptotic process by cytochrome c, positive regulation of mitochondrial outer membrane permeabilization involved in apoptotic signaling pathway, and apoptotic process involved in blood vessel morphogenesis. It has also been observed experimentally that BMP2 signaling modulates hypoxia-induced apoptosis, and this reduces PH severity in rats [112].

However, perhaps the most beneficial effects of BMP signaling are stimulating the expression of BMPR2 forming a beneficial feedback mechanism that prevents excessive vascular cell proliferation. BMP2 inhibits PAEC proliferation by reducing SRC activity [113], inhibiting the transcription of ERK1 and ERK2 [114], and activating the transcription of ID1 and ID3 [115] (Supplementary section 1.3.2 and Figure 5c, 5f).

Additionally, we performed a semantic radiate analysis with BMP2 as the source resulting in a significant association with bone mineralization and positive regulation of osteoblast proliferation (Table 1 row 18). Not all the effects of BMP2 signaling are beneficial for PH patients, as BMP2 signaling can induce vascular calcification greatly diminishing the therapeutic potential of BMP2 in the treatment of PH [116, 117, 118].

We also explored the function of BMP9 (GDF2) using a semantic radiate analysis (Table 1 row 19) and found that BMP9 is significantly associated with negative regulation of EC proliferation and negative regulation of vascular EC migration. This agrees with the experimentally observed function of BMP9, which is to preserve EC quiescence [117, 95, 119]. However, BMPR2 loss in PAECs reverses the effect of BMP9 [118, 120].

### 3.4 The interaction between G protein, IL6, activin, and BMP signaling in PH

Collectively, our analyses point towards that S1P, L-Glu, 12(S)HETE, IL6, and activin A stimulate the activity of signaling pathways that converge on SRC, ERK1, AKT, and MLC to cause many of the pathophysiological phenomena that characterize PH.

#### 3.4.1 Normally S1P and STAT3 stimulate each other forming a positive feedback mechanism that preserves endothelial barrier function (Figure 3b, 3g, and 3h)

IL6 signaling leads to the activation of the transcription factor STAT3 (Figure 3g) that stimulates the expression of SPHK1 (Figure 3h), SPHK2, and SPNS2. The enzymes SPHK1 and SPHK2 catalyze the phosphorylation of sphingosine to produce S1P. SPNS2 is a cell membrane protein that forms a channel and allows the transport of S1P from the cytoplasm to the extracellular space. S1P binds S1PR1 resulting in the activation of SRC, ERK1, and ERK2 (Figure 3b). SRC directly activates STAT3 and indirectly upregulates EGR1 through ERK1 (Figure 3g). STAT3 and EGR1 increase the expression of SPHK1 stimulating the production of S1P in human lung microvascular endothelial cells (Figure 3h). This positive feedback mechanism between S1P and STAT3 preserves endothelial barrier function [121]. However, ERK1 can also phosphorylate STAT3 on serine 727, which inhibits STAT3 dimerization and activation by preventing the phosphorylation of tyrosine residues on STAT3 [122] (Figure 3g). This constitutes a negative feedback mechanism that likely prevents excessive S1P production. We surmise that this negative feedback mechanism is ineffective in PH patients due to the higher concentration of S1P present in their serum.

#### 3.4.2 The interaction between G-protein signaling and IL6 signaling: S1P, L-Glu, 12(S)HETE, and IL6 stimulate pulmonary vascular cell proliferation and increase PAEC barrier permeability (Figure 3g)

S1P also signals through S1PR3 resulting in SRC and ERK activation [123]126 (Figure 3g). Additionally, L-Glu signaling through GRM2 and GRM3 positively modulates SRC activity (Figure 3g, Supplementary section 1.1.3). Further, 12(S)-HETE binds 12HETER and stimulates SRC activity [63] (Figure 3g). Therefore, we propose that in PH patients S1PR3, GRM2, GRM3, and 12HETER signaling causes the positive feedback mechanism that exists between STAT3 and S1P to be overactive [121] (Section 3.4.1). Notably, IL6, S1PR3, GRM2, GRM3, and 12HETER signal in concert to stimulate vascular cell proliferation, disrupt endothelial barrier function, and promote vasoconstriction (Supplementary sections 1.1.2-1.1.4, 1.2.2.-1.2.4).

#### 3.4.3 In PH patients activin signaling and S1P signaling amplify each other through positive feedback mechanisms that lead to excessive PAEC and PASMC proliferation (Figure 3h, and 3i)

GPCRs can transactivate receptor tyrosine kinases and serine/threonine kinase receptors, greatly amplifying their effect on cell signaling [124]. For instance, S1PR3 can activate SMAD3 during the differentiation of cultured human foreskin fibroblasts into myofibroblasts [125]. Vascular fibroblasts and myofibroblasts are very closely related to PAECs and PASMCs through the endothelial-to-mesenchymal transition and therefore we propose that this activation of SMAD3 also occurs in PAECs (Figure 3i). SMAD3 then stimulates the transcription of SPHK1 (Figure 3h) and SPHK2 increasing the concentration of S1P, forming a positive feedback mechanism that promotes excessive vascular cell proliferation in PH patients [109]. endothelin (ET1) binds to the GPCR endothelin receptor A (EDNRA) leading to SMAD2 phosphorylation in bovine aortic endothelial cells [126, 127]. Further, SMAD3 stimulates the production of ET1 [98, 110] (Figure 3h). We propose the existence of a positive feedback mechanism between SMAD2 or SMAD3 and ET1 that promotes vasoconstriction. A molecular mechanism for the transactivation of T*β*RIs by GPCRs (Figure 3i) involves the sequential activation of RHOA, ROCK, and Integrin. Then, active integrin causes the large latent TGF*β* complex to activate T*β*RIs resulting in the phosphorylation of SMAD2 leading to increased PAEC and PASMC proliferation [128].

#### 3.4.4 STAT3 and SMAD3 inhibit each other’s transcription and activation to stimulate PAEC and PASMC proliferation in PH patients (Figure 3f-3h)

In PH patients activin A and IL6 are present at a higher concentration than in the healthy population (Figure 3g) [80, 81, 96]. Activin A causes the activation of SMAD3 and IL6 signaling leads to the activation of STAT3 (Figure 3g). The crosstalk between STAT3 and SMAD3 depends on the specific biological context. We hypothesize that the following pathways might also be active in the PAECs and PASMCs of PH patients [129]; In cultivated human lung adenocarcinoma cells STAT3 represses the transcription of SMAD3 in cooperation with SKI and SKIL preventing SMAD2, SMAD3, and SMAD4 from activating the transcription of SMAD3 [130] (Figure 3h). STAT3 [131] and SMAD3 [132] also inhibit the activity of SMAD3 by inducing the expression of SMAD7 (Figure 3h). Also, in an immortalized human keratinocyte line, STAT3 directly binds phosphorylated SMAD3 and prevents it from binding SMAD4 and regulating transcription [133] (Figure 3h). By inhibiting SMAD3, STAT3 might allow PAEC and PASMC proliferation in PH patients. Further, in cultured human hepatocellular carcinoma cells SMAD3 and SPTBN1 collaborate to inhibit the ATF3, and CREB2-mediated transcription of STAT3 [134] (Figure 3h). Interestingly SMAD2, SMAD3, and STAT3 collaborate to activate the expression of SNAI1, one of the main transcription factors involved in the endothelial-to-mesenchymal transition that causes EC to stop expressing VE-cadherin, PECAM1, and VEGFR2 and detach from the intimal layer [91, 129, 135]. This constitutes an additional molecular mechanism through which STAT3 erodes endothelial barrier function.

### 3.5 The mechanism of action of commonly used and novel drugs for the treatment of PH

In this section, we aim to investigate how the mechanisms of action of currently used and novel pharmacological agents target the signaling pathways explored in the previous sections.

#### 3.5.1 Prostacyclin agonists, originally introduced as vasodilators, also target inflammation and inhibit PASMC proliferation (Figure 3b, 3d, and 3e)

Prostacyclin (PGI2) is a cardioprotective hormone produced and secreted by ECs. The synthesis of PGI2 begins with the liberation of arachidonic acid (AA) from membrane phospholipids, mainly by Ca2+ dependent phospholipase A2 (PLA2) isoforms. Then, AA is transformed into prostaglandin H2 by cyclo-oxygenases (COX1, COX2). After that prostacyclin synthase catalyzes the reaction that converts prostaglandin H2 into PGI2 [136] (Figure 3b).

PGI2 activates the Gs–coupled receptor IP and PPAR*β*. IP stimulates adenylate cyclase leading to an increase in cAMP [136, 137, 138], which activates PKA and EPAC1 [66]—leading to the deactivation of MLC, preventing vasoconstriction, and enhancing endothelial barrier function (Supplementary sections 1.1.2.-1.1.3). Further, PGI2 inhibits PASMC proliferation using a cAMP-dependent mechanism [139]. Specifically, cAMP sequentially activates PKA and CSK to inhibit SRC and prevent excessive PASMC proliferation [69]. PPAR*β* mediates many of the anti-inflammatory effects of PGI2. For instance, PPAR*β* activity inhibits TGF*β*-mediated IL6 upregulation [140] (Figure 3b, 3e).

The PGI2 agonists epoprostenol, iloprost, and treprostinil activate both IP and PPAR-mediated pathways, selexipag only activates IP receptors [141] (Figure 3d). The therapeutic potential of PGI2-agonists for the treatment of PH is substantial due to their vasodilatory, antiproliferative, and anti-inflammatory effects [142], suggesting that they should be included more often as part of the initial treatment. The ability of PGI2-agonists to inhibit IL6 is particularly promising. The concentration of IL6 is elevated in the serum of PH patients and is correlated with disease severity and risk of death [80, 81]. Additionally, IL6 overexpression causes PH [82]. Therefore, pharmacological interventions that diminish IL6 signaling may have therapeutic potential in some PH patients [81]. Although the IL6R inhibitor tocilizumab showed no benefit in a Phase 2 trial [143], other approaches including targeting IL6ST [144] and PGI2-agonists may result in more efficient PH treatment options that target IL6 signaling [145].

#### 3.5.2 PDE5 and ET1 inhibitors inhibit vasoconstriction by preventing MLC activation (Figure 3b, 3d,3e,3h, and 3j)

The PDE5 inhibitors sildenafil [146, 147] and tadalafil [148] were initially developed to treat erectile dysfunction. By inhibiting PDE5A1, sildenafil and tadalafil prevent the transformation of cGMP into 5’-GMP. Increased levels of cGMP then activate PKG I which prevents vasoconstriction by inhibiting PI3KR and averting the release of Ca2+ from the sarcoplasmic reticulum. Additionally, PKG I activates MLCP, which dephosphorylates MLC and hinders vasoconstriction [149, 150]. Both drugs decrease pulmonary vascular resistance and improve exercise capacity [146, 148](Figure 3b, 3e).

Another common approach to reduce vasoconstriction is to inhibit ET1 signaling. The expression of ET1 in the PAECs of PH patients is elevated compared to healthy individuals [151], possibly because SMAD3 upregulates the transcription of EDN1 which encodes ET1 [98, 110] (Figure 5g, 3h). Another relevant mechanism that could increase the expression of EDN1, involves the FOS and JUN that are upregulated by ERK signaling [74, 152] and stimulate the transcription of EDN1 [153, 154] (Figure 3j). All ET1 receptors belong to the GPCR superfamily of receptors.

PASMCs express two kinds of ET1 receptors; EDNRA is present at a higher concentration and mediates vasoconstriction, EDNRB is present at a lower concentration. In PASMCs, stimulation of both receptors by ET1 causes vasoconstriction [155, 156]. ET1-stimulated EDNRA can activate G12, G13, and Gq signaling [157, 158] (Figure 3i) causing vasoconstriction [159]164 (Supplementary section 1.1.2). Additionally, EDNRA signaling stimulates human PASMC proliferation [160] through ERK signaling [161]. This pathway is likely mediated by Gi/0 signaling [157, 158]. EDNRB also activates Gq increasing the concentration of Ca2+ in the cytosol through PLC*β* signaling leading to MLC activation and PASMC contraction (Figure 3b).

In PAECs the EDNRB-mediated increase in Ca2+ activates TRPV4, which is a transient receptor potential channel permeable to Ca2+, activates eNOS stimulating the production of NO [162] (Figure 3b). Additionally, Ca2+ activates PLA2, which releases AA from membrane phospholipids resulting in increased PGI2 production [136] (Figure 3b). The increase in NO and PGI2 causes vasodilation. EDNRB also functions as an ET1 trap and decreases its availability for EDNRA [163]. Several PH drugs target endothelin signaling (Figure 3d): bosentan [164, 165, 166] and macientan [167] are inhibitors of EDNRA and ENDRB that improve exercise endurance, hemodynamics, and survival in PH patients [167]. Other drugs like ambrisentan [168] selectively target EDNRA aiming to preserve ENDRB-mediated production of NO and PGI2 and allowing EDNRB to clear ET1 from circulation. However, this effect is very dependent on specific pathophysiological conditions that are largely unknown [169]. Selectively targeting EDNRA decreases the risk of increased liver aminotansferases and increases the risk of edema [169].

#### 3.5.3 Pharmacological approaches to restore the balance between activin and BMP signaling (Figure 3a, 3b,3c, 3f, 3g,3h,3i)

In PH patients activin signaling is upregulated and BMP2 signaling is downregulated [94, 95, 96, 98]. In PH patients the concentration of activin A and activin B is elevated [96, 98]. These ligands bind BMPR2 with a higher affinity than BMP2 and BMP4. Therefore, activins A and B downregulate BMP2 and BMP4 signaling through competitive inhibition [170] (Figure 3f). Additionally, activin signaling promotes the internalization and the proteasomal degradation of BMPR2 [96] (Figure 3g). Further, the expression of BMPR2 and ALK3 is markedly reduced in the pulmonary vasculature of PH patients [171]. This disbalance promotes excessive PASMC proliferation, vasoconstriction, and loss of PAEC identity resulting in perturbed endothelial barrier function. There have been three main approaches to restoring this balance.

The canonical effectors of activin signaling are SMAD2 and SMAD3, simvastatin activates PP2A and PPM1A that dephosphorylate SMAD2 and SMAD3 [172]. However, in a phase 1 trial, simvastatin only produced a small and transient reduction in RV mass [173]. Based on our analysis the inefficacy of simvastatin as a PH treatment might be because SMAD-independent activin signaling promotes vasoconstriction and PASMC proliferation (Section 3.4.1), also because PPM1A inhibits SMAD1, SMAD5, and SMAD9 activity (Figure 3c), which can prevent excessive vascular cell proliferation (Section 3.4.2).

Tacrolimus (FK506), which inhibits calcineurin and is used to prevent transplanted organ rejection, released FKBP12 from ALK1, ALK2, and ALK3 allowing BMP4 to activate SMAD1 and SMAD5 leading to enhanced BMPR2 signaling in cultured PAECs from PH patients [174] (Figure 3i). A phase IIa trial of tacrolimus as a PH treatment concluded that it had beneficial effects on 3 end-stage patients [175]. However, another phase IIa trial found that the beneficial effects of tacrolimus were not significant [176]. Increasing BMP4 signaling in PASMCs leads to their proliferation and upregulates vasoconstriction [112]. Further, calcineurin is involved in a signaling pathway that also involves S1P, PLC, NFATC3, and OPN that leads to PASMCs proliferation [177]. By upregulating BMP4 signaling and inhibiting calcineurin, we surmise that tacrolimus has an unpredictable effect on PASMC proliferation that depends on the pathophysiological context of each patient.

Sotatercept, which was originally developed for the treatment of anemia and bone loss [178], is under trial as a promising treatment for PH [97, 179]. Sotatercept is a soluble ACVR2A IgG-Fc fusion protein that functions as a trap for activin A, activin B, BMP9, GDF8, GDF11 and other ligands from the TGF*β* family that have a high binding affinity for ACVR2A [170] (Figure 3a). Sotatercept increases BMPR2 availability by preventing activin binding and BMPR2 degradation (Figure 3f. 3g), this allows BMP2 to activate SMAD1, SMAD5, and SMAD9 inhibiting the transcription of ERK1 and ERK2 [114], and activating the transcription of BMPR2 [117], ID1 and ID3 [115] (Figure 3c). Additionally, when BMP2 binds BMPR2 it decreases SRC phosphorylation in PASMCs [113] (Figure 3b). The effect of these BMP2-associated pathways is to prevent excessive vascular cell proliferation, preserve PAEC barrier function and prevent vasoconstriction. Downregulating activin signaling decreases SMAD2 and SMAD3 activity, and this prevents excessive S1P and ET1 production (Figure 3h). Additionally, downregulating activin signaling prevents SMAD-independent activation of SRC and ERK1 (Figure 3g) inhibiting GPCR-mediated vasoconstriction, endothelial barrier disruption and PASMC proliferation. In summary, sotatercept prevents the excessive activation of SRC, ERK1, AKT, and MLC, the molecules we identified as convergence nodes in the interplay between different PH signaling pathways.

#### 3.5.4 Seralutinib prevents excessive PASMC by inhibiting PDGF signaling (Figure 3i, 3j)

PDGF signaling is one of the main molecular mechanisms that induce PASMC proliferation. PAECs, pulmonary vascular pericytes, and PASMCs express PDGFA, PDGFB, PDGFR*α* and PDGFR*β* and these genes are overexpressed in the lung arteries of PH patients [180, 181]. In PASMCs, PDGFB signaling induces ERK1-mediated cell proliferation and stimulates cell survival and growth through AKT1, and mTOR [180] (Figure 3i). In rat PASMCs, PDGFB-stimulated ERK1 and AKT signaling and upregulates the expression of SPHK1 and the production of S1P promoting PASMC proliferation [182]. In mesangial cells that are contractile, and perivascular like PASMCs, JUN, and FOS that are upregulated by ERK signaling, activate the transcription of SPHK1 [183] (Figure 3j). Additionally, S1P signaling increases the expression of miR-21, which decreases the expression of BMPR2 and ID1 allowing excessive PASMC proliferation [182, 115]. Also, *β*-catenine (BC), which promotes PASMC proliferation, is elevated in PH patients. Moreover, PDGFB signaling prevents GSK3*β*-mediated BC degradation and promotes BC nuclear localization and transcriptional activity in PASMCs. Further, BC knockdown reduces PDGFB-stimulated PASMC proliferation [184]. Lastly, from a metabolic perspective, it is interesting to note that in PASMCs PDGFB, AKT1, mTOR, and HIF1A signaling is associated with the Warburg effect [185], defined as an increase in glucose breakdown and lactate production even when sufficient oxygen is available. The Warburg effect may cause excessive PAEC and PASMC proliferation in PAH patients [186].

Inhibiting PDGFR*α*, PDGFR*β*, and other tyrosine kinases systemically using imatinib leads to reduced PASMC proliferation. However, it is associated with very serious side effects including an increased risk of subdural hematoma preventing its use in the treatment of PH [187]. Seralutinib is a novel tyrosine kinase inhibitor delivered via inhalation that downregulates PDGFR*α*, and PDGFR*β* signaling specifically in the lungs (Figure 3i). Seralutinib is currently under phase 2 clinical trial [188] and its effect on rats and PH lung samples was promising and restored the pulmonary expression of BMPR2 [189]. All the hypotheses generated by our analysis are summarized in tables 2 and 3.

**Table 2:**
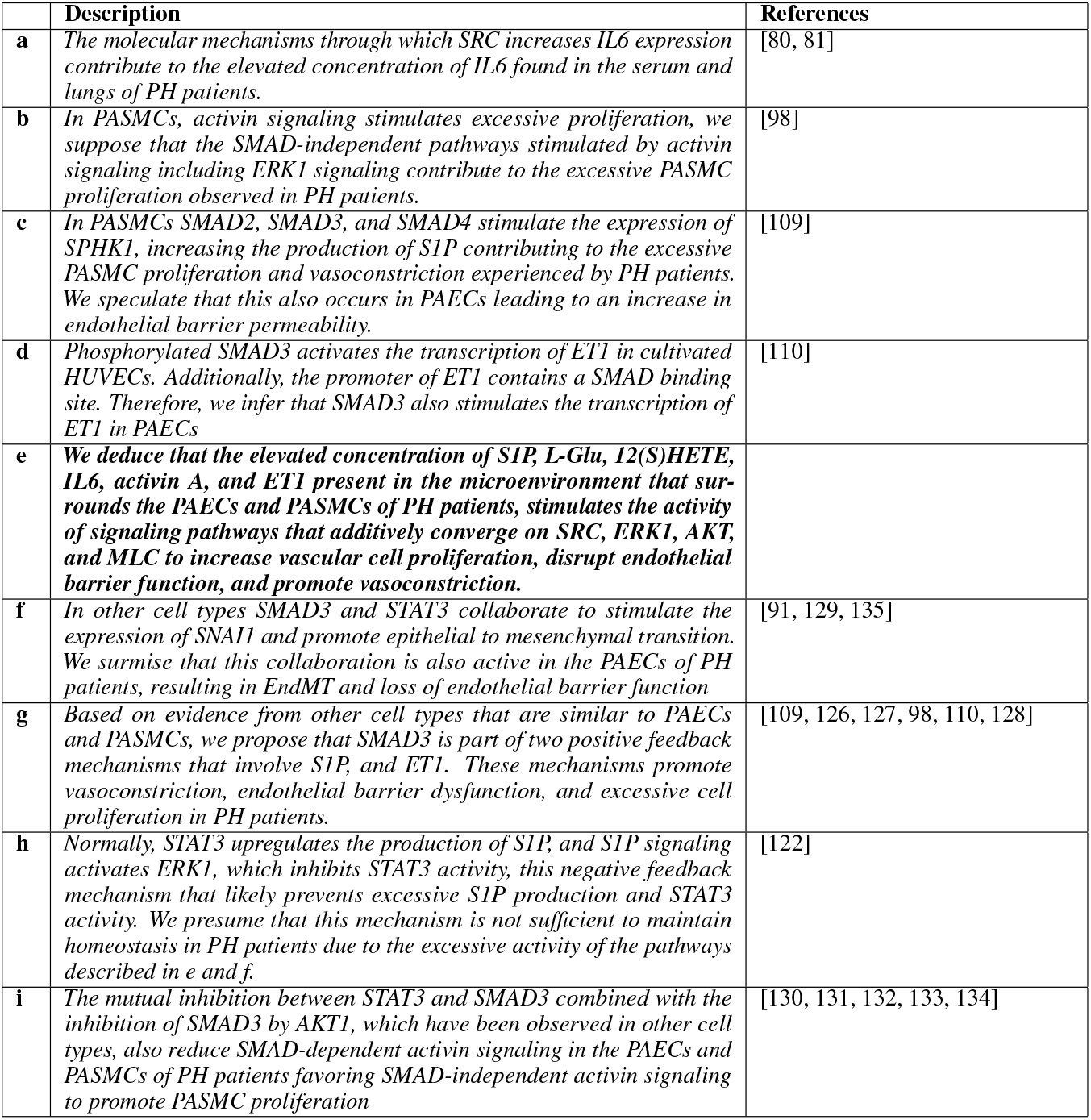
Hypotheses about physiological mechanisms.

**Table 3:**
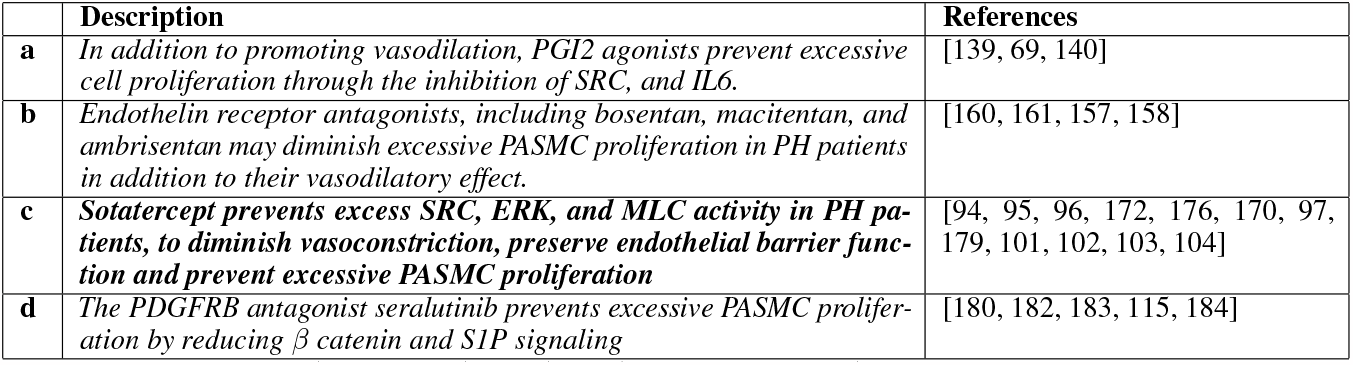
Hypotheses about pharmacological mechanisms of action.

## 4 Discussion

Despite substantial progress in the understanding of PH pathophysiology over the last years, the detailed mechanisms underlying PH remain elusive due to the complex interplay of dysregulated biological processes. Also, although the mechanisms of action of some drugs that have been successfully used for the treatment of PH have been proposed, certain drugs may show additional mechanisms of action that are yet to be identified. Understanding these additional mechanisms of action is crucial for the personalized and thus more effective treatment of PH patients. In this study, we aimed to generate detailed mechanistic hypotheses of pathophysiological processes in PH using our in-house developed software platform *Lifelike*. Importantly, we have focused our analyses on the potential effects of blood plasma metabolites from PH patients on pulmonary endothelial and smooth muscle cells. Moreover, we used *Lifelike* to investigate the mechanisms of action of existing and new drugs used for the treatment of PH in more detail.

### 4.1 Integrating information from several sources using *Lifelike* enabled us to generate novel hypotheses about potential molecular mechanisms involved in the pathophysiology of PH

Using only the four differentially abundant blood plasma metabolites S1P, 12(S)HETE, L-Glu, and L-Arg of PH patients as a starting point, this study demonstrates how knowledge mining with *Lifelike* enabled us to:

1. generate detailed mechanistic hypotheses about PH pathophysiology,
2. gain insights into the complex interplay of signaling pathways relevant to PH,
3. suggest mechanisms of drug actions in addition to those previously reported (e.g. sotatercept) that might impact personalized treatment of PH patients in the future, and
4. capture new knowledge effectively by creating *Lifelike* knowledge maps with links to references.

We have achieved this by analyzing metabolite data and receptor ligands on the curated database Reactome using graph algorithms, and by combining our software’s additional data analytics functionalities, including statistical enrichment and semantic analysis of textual information from *Lifelike*’s knowledge graph. Additionally, we validated, interpreted, and extended our results based on relevant existing literature. This entire process allowed us to explore the pathophysiology of PH and to propose the model shown in figure 3 and summarized in figure 6. Further, *Lifelike* allowed us to generate the hypotheses discussed in the following sections and shown in Tables 2 and 3.

**Figure 6:**
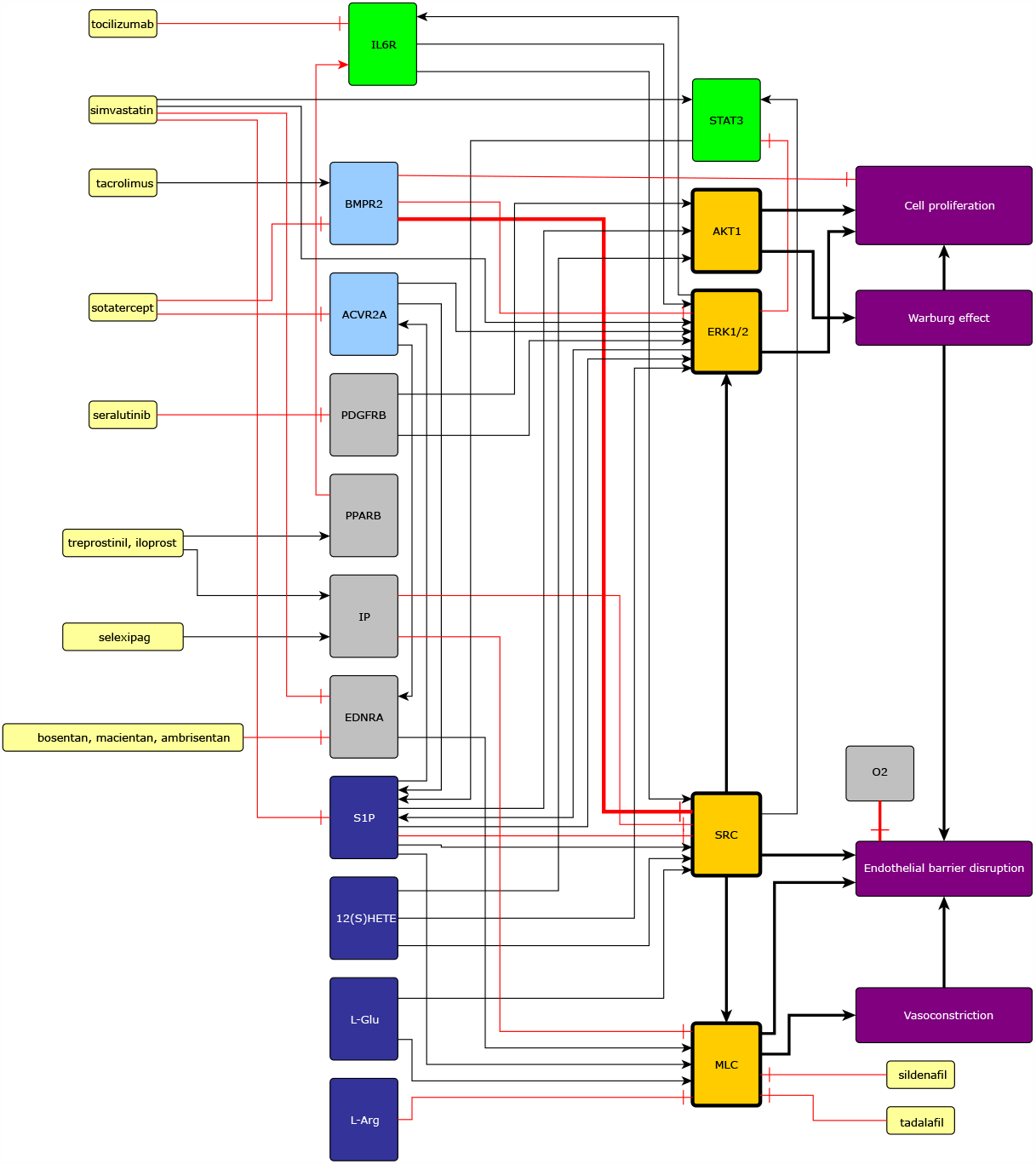
Currently available pharmaceutical treatments, GPCRs, IL6, actvin, BMP, PPARB, and PDGFB signaling converges on SRC, MLC, ERK1, and AKT. Leading to PAEC and PASMC proliferation, endothelial barrier dysfunction, vasoconstriction, and the Warburg effect. Black arrows represent activation, and red arrows represent inhibitions. Yellow nodes are medicines, green nodes are part of the IL6 signaling pathway, light blue nodes are part of the activin or BMP signaling pathways, convergence nodes are shown in orange, dark blue nodes are differentially expressed metabolites, and purple nodes represent physiological effects. This figure also contains potential mechanisms of drug action proposed in this study.

*Lifelike* also enables the reader of a biology article to quickly find the molecules, diseases, and phenotypes that are discussed in the manuscript or included in the figures through entity recognition. For instance, genes detected in articles by *Lifelike’s* entity recognition system can be used to create an enrichment table where the genes can be associated with biological processes, molecular functions, and cell structures by GO enrichment. This allows the reader to find context-specific information quickly, and to extend the knowledge beyond what is described in the article with other sources of information (Table 1 row 21) We encourage the reader to upload this manuscript in PDF format into *Lifelike* for improved reading and knowledge extraction capabilities. *Lifelike* knowledge maps aid in the development of custom high-quality knowledge reconstructions with links to relevant literature through manual curation (Table 1 row 22). We envision developing the computational tools that will enable us to transform these curated networks into graph databases that can be then integrated with relevant existing databases or used on their own for graph analysis.

### 4.2 Interaction between different signaling pathways (Table 2b-2f)

The prevailing reductionist research paradigm focuses on one molecule, or at most one pathway at a time [190]. Currently, the presence and activity of many molecules can be experimentally measured simultaneously. Additionally, many molecules and pathways have been carefully analysed and deposited in curated databases. This allows us to explore how molecular pathways influence each other, forming complex dynamic systems characterized by emergent properties that cannot be explained by a single molecule, pathway, or cell [190, 191, 192] Finding these emergent properties to develop a holistic understanding of a disease for the improvement of current treatments and future drug development is the goal of many research efforts in medical systems biology [193] including the present study. The convergence of several pathways on certain molecules is an important emergent property of the network model presented in figure 3. This convergence allowed us to generate several hypotheses (Table 2b-2f). Notably, the elevated concentration of S1P, L-Glu, 12(S)HETE, IL6, activin A, and ET1 present in the microenvironment that surrounds the PAECs and PASMCs of PH patients, stimulates the activity of signaling pathways that additively converge on SRC, ERK1, AKT1, and MLC to increase vascular cell proliferation, disrupt endothelial barrier function, and promote vasoconstriction (Figure 6). AKT1 is associated with cell proliferation and the Warburg effect which promotes cell proliferation and endothelial barrier dysfunction. SRC activates MLC and ERK1 and causes endothelial barrier dysfunction. ERK1 induces cell proliferation. MLC activity leads to vasoconstriction and endothelial barrier dysfunction (Figure 6).

### 4.3 Potentially important feedback mechanisms (Table 2g-2i)

In dynamic systems, positive feedback mechanisms that contain an even number of inhibitory edges, are associated with multistability, which can be interpreted as phenotypic diversity in biological systems [194, 195, 196]. Alternatively, positive feedback mechanisms can amplify signals [197]. Based on evidence from other cell types that share many important characteristics with PAECs and PASMCs, we propose that SMAD3 is part of two positive feedback mechanisms that involve S1P (Figure 7b), and ET1 (Figure 7a). These mechanisms promote vasoconstriction, endothelial barrier dysfunction, and excessive cell proliferation in PH patients.

**Figure 7:**
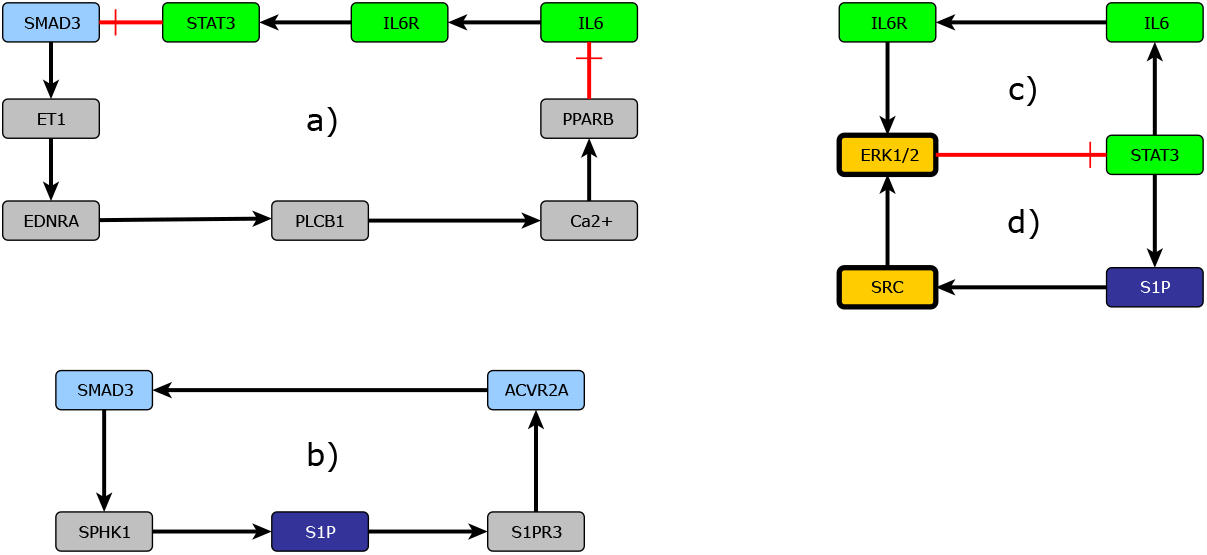
Feedback mechanisms: a) A positive feedback mechanism between SMAD3 and ET1, b) A positive feedback mechanism that involves S1P and SMAD3, c) A negative feedback mechanism that involves IL6, d) A negative feedback mechanism that involves SP1. Black arrows represent activation, and red arrows represent inhibitions. Green nodes are part of the IL6 signaling pathway, light blue nodes are part of the activin or BMP signaling pathways, convergence nodes are shown in orange, and dark blue nodes are differentially expressed metabolites,

Negative feedback mechanisms, characterized by the presence of an odd number of inhibitory edges, may cause or allow cyclic behavior, like the cell cycle and the circadian rhythm [195, 196, 198, 199]. Alternatively, negative feedback circuits can balance the activity of two molecules or pathways, in biological systems this can contribute to homeostasis and robustness [200]. The intrinsic robustness against perturbations of biological systems regulated by molecular regulatory networks and morphogen gradients might be one of the main causes that prevent pharmacological compounds from having the desired therapeutic effect [201]. The role of robustness in the effectiveness of PH treatment remains largely unknown. Normally, S1P is present at a concentration that is only sufficient to activate S1PR1 signaling to preserve PAEC barrier function, and the regular activity of IL6 signaling prevents PAEC apoptosis and hypoxia-induced PH. Two negative feedback mechanisms might prevent the excessive IL6 and S1P activity observed in PH patients and maintain homeostasis. S1P signaling induces SRC that activates ERK1, which inhibits STAT3 activation preventing an increase in the production of S1P (Figure 7d). Similarly, IL6 signaling also activates ERK1, inhibiting the activation of STAT3 and the subsequent transcription of IL6 (Figure 7c).

### 4.4 Hypotheses concerning the mechanism of action of existing and novel pharmacological treatments of PH (Table 3) and their clinical implications

PH is a complex disease process involving multiple molecular mechanisms and chronic environmental, metabolic, genetic, and hypoxic insults resulting in pathologic remodeling of primarily medium- and small-sized pulmonary arterioles [202]. The progression of pulmonary vascular remodeling in PH is driven by dynamic and local dysfunctions of both the pulmonary endothelium and vascular smooth muscle. In addition, abnormalities that affect the inflammatory and immune compartment is essential to produce the disease phenotype [203] Conventional therapies of currently approved PH drugs, all target vasoconstriction [203]. Available PH medications include endothelin receptor antagonists (ERAs), phosphodiesterase type-5 inhibitors (PDE-5i), soluble guanylate cyclase stimulators, prostacyclin analogs, and oral prostacyclin receptor agonists none of which are curative [1]212. Using *Lifelike*’s ability to find novel potentially important drug mechanisms of action, enables pharmacological repurposing by finding additional mechanisms of action for existing drugs or proposing the potential mechanism of action of novel drugs. We were able to associate existing and novel drugs with molecular mechanisms of action that affect endothelial barrier dysfunction and PASMC proliferation.

The quest for specific molecules that can influence signal transduction has become an essential part of exploring potential new drugs for the treatment of PH. In PH signaling pathways are stimulated that converge on SRC, ERK, AKT, and MLC leading to excessive vasoconstriction, PASMC proliferation, and endothelial barrier dysfunction. The mechanisms of action of the existing and novel treatments for PH also converge on these 4 molecules (Figure 6). ERAs and PGI2 agonists are used mostly due to their known vasodilatory effect, our work emphasizes that they also prevent excessive cell proliferation through the inhibition of SRC. The PDGFRB antagonist seralutinib prevents excessive PASMC proliferation by downregulating *β* catenin and S1P signaling. *Lifelike* allowed us to understand that sotatercept is exceptional because it simultaneously targets SRC, ERK, AKT, and MLC by modulating SMAD-independent activin and BMP signaling, not mentioned in previous descriptions of the mechanism of action of sotatercept [178, 97].

### 4.5 Limitations of our methodology and future perspectives

When Arturo Rosenblueth and Norbert Wiener discussed the role of models in biology, they proposed that “The best material model for a cat is another or preferably the same cat”. However, for practical reasons, it is necessary to reduce the complexity of models until it is possible to analyze and understand them [204]. PH is a complex disease that involves vasoconstriction, endothelial barrier function, vascular cell proliferation, and other important biological mechanisms including the molecules that regulate oxidative stress, coagulation, inflammation, mechanosensory mechanisms, and EC cytoskeleton remodeling, many of which are not included in our model. Moreover, the genetic mutations and the comorbidities that afflict each specific PH patient can affect the molecular mechanisms involved in PH. Additionally, from a cellular perspective, we focused on PAECs and PASMCs. Macrophages, platelets, fibroblasts, and other cell types are also important in PH. Further, PH leads to right ventricle hypertrophy and other pathological processes that occur in other organs. It would be interesting to include these additional biological mechanisms, cell types, and organs in a future version of our model. The effectiveness of graph algorithms is highly dependent on the accuracy, completeness, and quality of the knowledge graph. Incomplete or inaccurate graphs can lead to misleading results. Path-finding algorithms often prioritize the shortest path, which might not always represent the most meaningful or relevant connection between two nodes. Centrality measures might sometimes overly prioritize nodes that are merely popular or have more connections. Incorporating additional contextual data, like gene expression datasets or gene mutations, can guide path-finding algorithms more effectively. If the knowledge graph is not regularly updated with fresh information, it will become outdated, leading to analyses that no longer reflect the current state of knowledge or understanding in a particular field. Moving forward, we aim to employ graph embeddings to anticipate new connections between nodes, fostering the generation of novel hypotheses. Another important limitation of our methodology is that certain interactions between molecules and their effect have been observed only in other cell types, model organisms, or cells cultivated under conditions that differ from those encountered in vivo. Also, we assembled a topological model of the network of molecules involved in PH. These molecules constitute a dynamic system. Using a mathematical formalism like a Boolean network, a Petri net, or a set of ordinary differential equations would allow us to simulate and explore the behavior of this dynamic system including the effect of the different existing and novel pharmacological treatments. A mathematical formalism would also allow us to find important missing connections in the network [205]. Lastly, modeling in general, including this study only allows us to summarize, analyze, and propose novel hypotheses. Importantly all the hypotheses we generated require experimental validation.

## 5 Conclusions

Pulmonary hypertension (PH) constitutes a complex disease process affecting 1% of the global population. Despite major advances in the treatment, PH remains a devastating disease involving multiple pathophysiological mechanisms including genetic, molecular, and environmental factors. There is a pressing need for more potent, disease-modifying clinical therapies to enhance long-term outcomes for PH patients. A comprehensive review of the PH signaling pathways involved in cell proliferation, endothelial barrier disruption, vasoconstriction, and the Warburg effect is presented using the software platform *Lifelike*. The molecular mechanisms we have elucidated using *Lifelik*e open a window for pharmacological repurposing. By identifying new actions for existing drugs and suggesting potential mechanisms for new drugs, we highlight new promising therapeutic targets for this devastating group of diseases.

## Supporting information

Enriching a Figure

Pulmonary hypertension journey

Pathways map

## Data Availability

All data used for this study can be found in the manuscript or at: https://doi.org/10.1038/s41598-022-17374-x and https://zenodo.org/doi/10.5281/zenodo.6109727

https://zenodo.org/doi/10.5281/zenodo.6109727

## Author Contributions

NW, JC, SS, HH, ET, and PJ planned the research, revised the manuscript for important intellectual content, analyzed, and discussed the results. NW, JC, ET, and SS reviewed the literature, NW and SS made the tables and the figures. NW, SS, and ET used *Lifelike* to research pulmonary hypertension. TS provided support on graph database creation and algorithms. ET, JC, and PJ obtained funding for this project.

## Acknowledgments

We thank Ethan Daniel Sanchez, Michael Siu, Dominik Mariusz Maszczyk, Robin Cai, Ewa Lis, David Regla Demaree, Sharon Wiback, Maciej Tomasz Grzelczak, and Christian Degnbol Madsen for the development and upkeep of *Lifelike*. We greatly appreciate the help of Rikke Stine Friis Fléron.

