## Supplementary material for "A *Lifelike* guided journey through the pathophysiology of pulmonary hypertension - from measured metabolites to the mechanism of action of drugs": Enriching a Figure

1) Upload and generate annotations

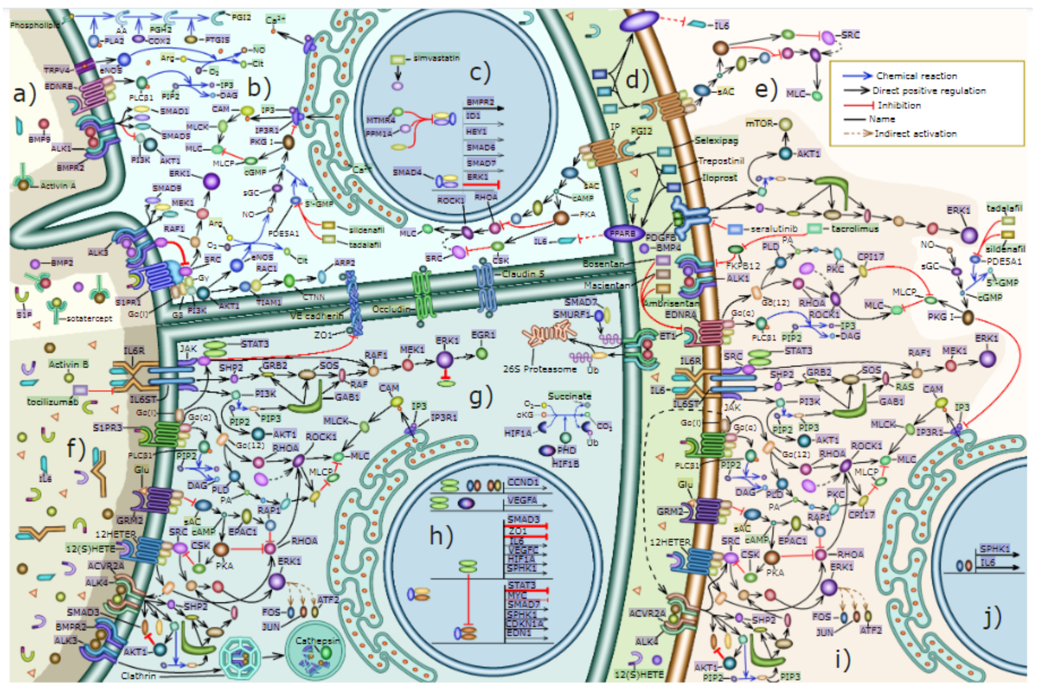

2) Explore the annotations using word clouds

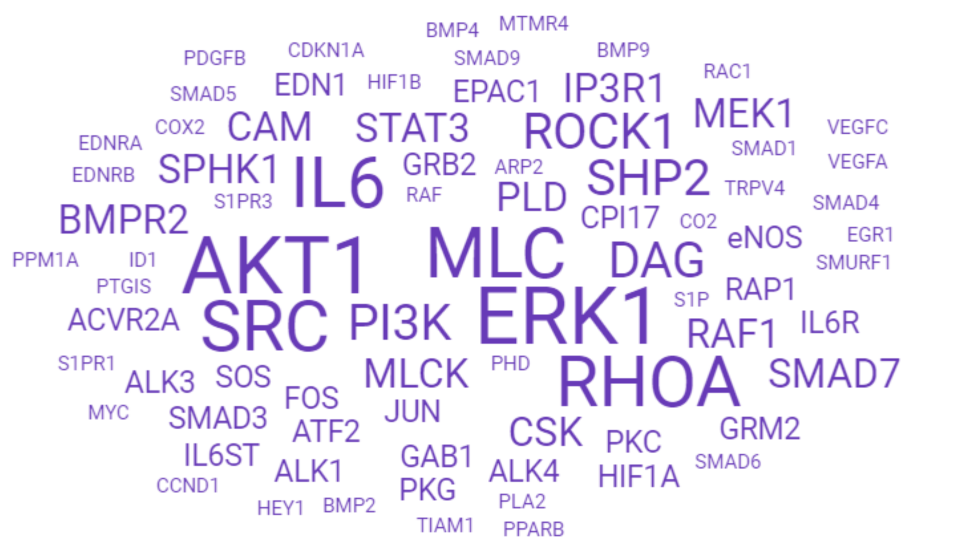


Genes found in the figure

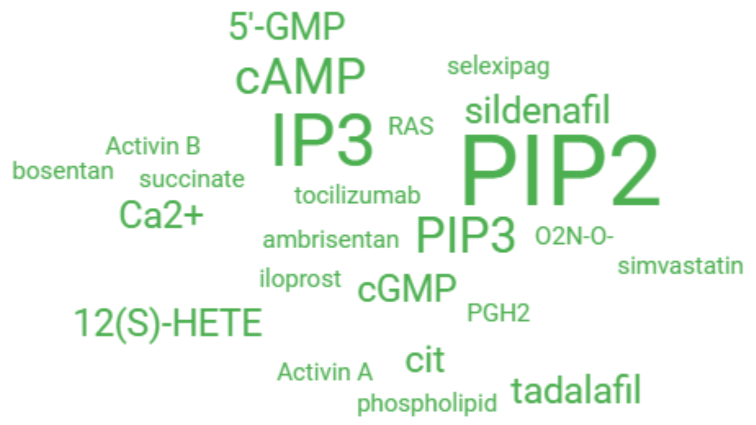


Chemicals that appear on the figure

3) Use the genes to create an enrichment table

interaction
figure genes

4) Find relevant biological processes through statistical enrichment.

Biological Process — Table | Graph | Clustergram | Cloud

Search:

| GOID | GO Term | Associated genes | q-value |
|---|---|---|---|
| 30509 | **BMP signaling pathway** | BMPR2, ACVRL1, SMAD4, ACVR2A, SMAD6, SMAD3, MAPK3, SMURF1, BMP2, SMAD1, ID1, BMPR1A, GDF2, EGR1, SMAD5, SMAD7, SMAD9, BMP4 | 8.91e-21*** |
| 7179 | **transforming growth factor beta receptor signaling pathway** | RHOA, ACVRL1, SMAD4, SRC, MTMR4, SMAD3, SMAD6, JUN, SMURF1, SMAD1, ID1, BMPR1A, FOS, SMAD5, SMAD7, SMAD9 | 1.19e-15*** |

This tells us that BMP signaling is important.

Biological Process — Table | Graph | Clustergram | Cloud

Search: endothelial

| GOID | GO Term | Associated genes | q-value |
|---|---|---|---|
| 1938 | **positive regulation of endothelial cell proliferation** | ARNT, HIF1A, BMPR2, ACVRL1, AKT1, PDGFB, JUN, BMP2, GDF2, VEGFC, VEGFA, PIK3CD, BMP4 | 1.27e-13*** |
| 10595 | **positive regulation of endothelial cell migration** | BMPR2, EDN1, AKT1, PIK3CG, PIK3CB, GPLD1, RAC1, VEGFA, PIK3CD, BMP4 | 1.09e-9 *** |
| 48010 | **vascular endothelial growth factor receptor signaling pathway** | RHOA, ROCK1, SRC, PIK3CB, RAC1, VEGFC, VEGFA, PIK3CA | 5.69e-7 *** |
| 43536 | **positive regulation of blood vessel endothelial cell migration** | HIF1A, NOS3, AKT1, PDGFB, VEGFC, VEGFA | 3.60e-5 *** |
| 10575 | **positive regulation of vascular endothelial growth factor production** | ARNT, HIF1A, PTGS2, IL6, IL6ST | 5.55e-5 *** |

Endothelial cell proliferation and migration are affected by these genes.

Biological Process — Table | Graph | Clustergram | Cloud

Search: smooth muscle cell

| GOID | GO Term | Associated genes | q-value |
|---|---|---|---|
| 48661 | **positive regulation of smooth muscle cell proliferation** | EDN1, AKT1, PTGS2, IL6R, S1PR1, IL6, PDGFB, BMP4 | 2.73e-7*** |
| 1904707 | **positive regulation of vascular associated smooth muscle cell proliferation** | JUN, EDN1, PDGFB, BMPR1A | 7.55e-3** |

These genes promote smooth muscle cell proliferation.

Biological Process — Table | Graph | Clustergram | Cloud

Search: pressure

| GOID | GO Term | Associated genes | q-value |
|---|---|---|---|
| 3100 | **regulation of systemic arterial blood pressure by endothelin** | RHOA, NOS3, EDN1 | 3.62e-4*** |
| 8217 | **regulation of blood pressure** | ACVRL1, NOS3, EDNRA, PTGS2, EDNRB | 2.36e-3** |

Processes associated with blood pressure control

5) The word cloud asssciiatedd with the enrichment  table also contains valuable information

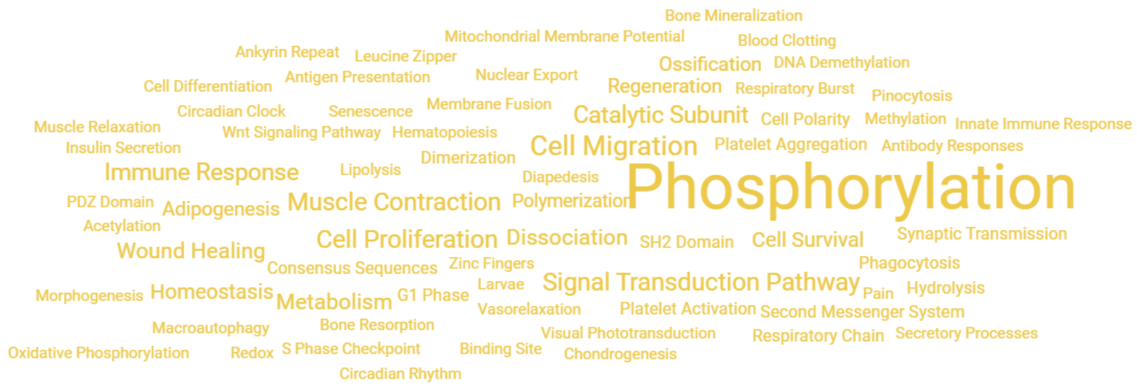


The phenomena associated with the genes on the enrichment table

After Phosphorylation, Signal Transduction Pathway, Cell Migration, Muscle Contraction, Cell Proliferation, and Immune Response,  stand out. If we look closely, we can also see Vasoconstriction, Blood Clotting, Platelet Activation, Ossification, and G1 phase. All these phenomena are important for pulmonary hypertension.

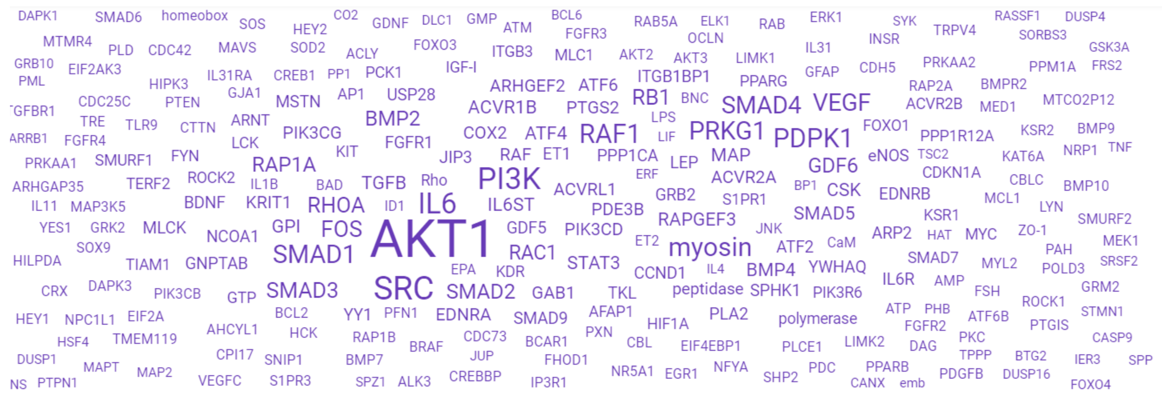


Genes present in the description of the enrichment table.

6) The genes names present in the description of the genes  can be used to create a larger enrichment table and  reach additional information.

interaction(1
)

Created by Lifelike
lifelike.bio
