## Supplementary material for "A *Lifelike* guided journey through the pathophysiology of pulmonary hypertension - from measured metabolites to the mechanism of action of drugs": Pulmonary hypertension journey

### 1) Radiate analysis with the upregulated and downregulated metabolites as sources

Radiate_analysis_for_s1p_glu_12hete_hyp_arg_hist.xlsx

| | |
|---|---|
| R-ALL-83957 | Hyp [extracellular region] |
| R-ALL-351977 | L-Arg [extracellular region] |
| R-ALL-210382 | L-Glu [extracellular region] |
| R-ALL-419305 | S1P [extracellular region] |
| R-ALL-3296298 | 12(S)-HETE [extracellular region] |
| R-ALL-351970 | L-His [extracellular region] |

Sources

| stId | displayName | pagerank |
|---|---|---|
| R-HSA-380091 | Ligand:GPCR complexes that activate Gi [plasma membrane] | 0,042766791 |
| R-ALL-179747 | ligands of SLC29A2 [extracellular region] | 0,039404389 |
| R-HSA-109527 | Equilibrative transport (import) of nucleosides and free bases by solute carrie | 0,031465133 |
| R-ALL-179742 | ligands of SLC29A2 [cytosol] | 0,029183131 |
| R-HSA-3499321 | GPR31:12(S)-HETE [plasma membrane] | 0,028349993 |
| R-HSA-749445 | Ligand:GPCR complexes that activate Gi:Heterotrimeric G-protein Gi (active) [ | 0,027604233 |
| R-HSA-109529 | Equilibrative transport (export) of nucleosides and free bases by solute carrie | 0,024589483 |
| R-HSA-749454 | The Ligand:GPCR:Gi complex dissociates | 0,023472061 |
| R-HSA-749455 | Ligand:GPCR complexes that activate Gi:Heterotrimeric G-protein Gi (inactive) | 0,022660246 |
| R-HSA-380073 | Liganded Gi-activating GPCR acts as a GEF for Gi | 0,01782534 |
| R-HSA-109527 | Equilibrative transport (import) of nucleosides and free bases by solute carrie | 0,016225859 |
| R-HSA-3296304 | GPR31 is a receptor for 12(S)-HETE | 0,015324623 |
| R-ALL-113599 | Hyp [cytosol] | 0,014646529 |
| R-HSA-749456 | Liganded Gi-activating GPCRs bind inactive heterotrimeric G-protein Gi | 0,012247748 |
| R-HSA-9690450 | S1PR1:S1P [plasma membrane] | 0,011339998 |
| R-HSA-419346 | S1PR2-5:S1P [plasma membrane] | 0,011339998 |
| | guanyl-nucleotide exchange factor activity of Ligand:GPCR complexes that act | 0,009633262 |
| R-ALL-375468 | SLC6A14 ligands [extracellular region] | 0,009556406 |
| R-HSA-167434 | G-protein beta-gamma complex [plasma membrane] | 0,007987795 |
| R-HSA-392161 | G alpha (i): GTP [plasma membrane] | 0,007553266 |

The most significant targets include many components of G protein signaling pathways

Forward_traces_for_s1p_glu_12hete_hyp_arg_hist.graph

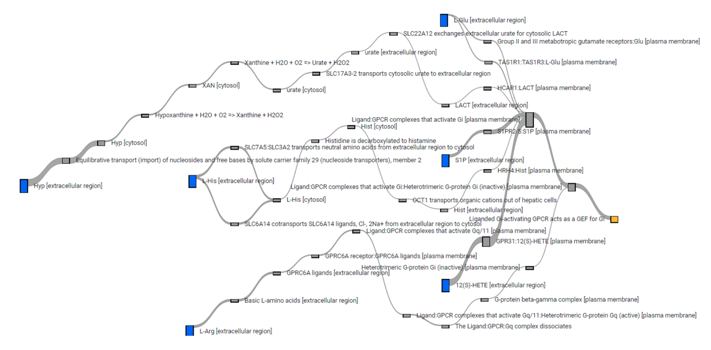

S1P, 12(s)hete, and L-Glu activate Gi signaling

### 2) SRC is a convergence point for S1P, 12(s)HETE ,and L-Glu

| stId | displayName | geneName | pagerank |
|---|---|---|---|
| R-HSA-8964276 | p-Y416-SRC [cytosol] | SRC | 0,000928258 |
| R-HSA-8964323 | Active BTK [cytosol] | BTK | 0,000172967 |
| R-HSA-391959 | P2RY12 [plasma membrane] | P2RY12 | 0,000127777 |
| R-HSA-195017 | ARHGEF6 [cytosol] | ARHGEF6 | 6,77634E-05 |
| R-HSA-8964622 | Active PAK1 [cytosol] | PAK1 | 6,77634E-05 |
| R-HSA-391837 | TBXA2R [plasma membrane] | TBXA2R | 4,71603E-05 |
| R-HSA-418898 | ADORA2B [plasma membrane] | ADORA2B | 2,6797E-05 |
| R-HSA-391956 | P2RY1 [plasma membrane] | P2RY1 | 2,39873E-05 |
| R-HSA-391834 | PTGIR [plasma membrane] | PTGIR | 2,3563E-05 |
| R-HSA-167679 | p-4Y-PLCG1 [plasma membrane] | PLCG1 | 1,83755E-05 |
| R-HSA-428935 | p-T180,Y182-MAPK14 [cytosol] | MAPK14 | 1,48092E-05 |
| R-HSA-194371 | p-T,Y-MAPK8 [cytosol] | MAPK8 | 1,02316E-05 |
| R-HSA-399816 | p-T508-LIMK1 [cytosol] | LIMK1 | 8,80459E-06 |
| R-HSA-186833 | RASA1 [cytosol] | RASA1 | 7,66306E-06 |
| R-HSA-162629 | PAK1 [cytosol] | PAK1 | 7,64617E-06 |
| R-HSA-210514 | STXBP1-1 [cytosol] | STXBP1 | 7,12242E-06 |
| R-HSA-168786 | SYT1 [plasma membrane] | SYT1 | 7,12242E-06 |
| R-HSA-210461 | RAB3A [plasma membrane] | RAB3A | 7,12242E-06 |
| R-HSA-210385 | CPLX1 [cytosol] | CPLX1 | 7,12242E-06 |
| R-HSA-195131 | SRGAP2 [cytosol] | SRGAP2 | 6,87986E-06 |
| R-HSA-1234131 | 2xHP-HIF1A [cytosol] | HIF1A | 5,95927E-06 |
| R-HSA-1234149 | 2xHP-EPAS1 [cytosol] | EPAS1 | 5,95927E-06 |

The non-receptor tyrosine knases SRC and BTK are the most significant protein targets of the metabolites


Watterson, K. R., Ratz, P. H. & Spiegel, S. The role of sphingosine-1-phosphate in smooth muscle contraction. Cell Signal 17, 289–298 (2005).

Murakami, A. et al. Sphingosine 1-Phosphate (S1P) Regulates Vascular Contraction via S1P3 Receptor: Investigation Based on a New S1P3 Receptor Antagonist. Mol Pharmacol 77, 704–713 (2010).

Tang, D. G. et al. The Lipoxygenase Metabolite, 12(S)-HETE, Induces a Protein Kinase C-Dependent Cytoskeletal Rearrangement and Retraction of Microvascular Endothelial Cells. Exp Cell Res 207, 361–375 (1993).

Liu, P. et al. Inhibition of Src activation reverses pulmonary vascular remodeling in experimental pulmonary arterial hypertension via Akt/mTOR/HIF-1<alpha> signaling pathway. Exp Cell Res 380, 36–46 (2019).


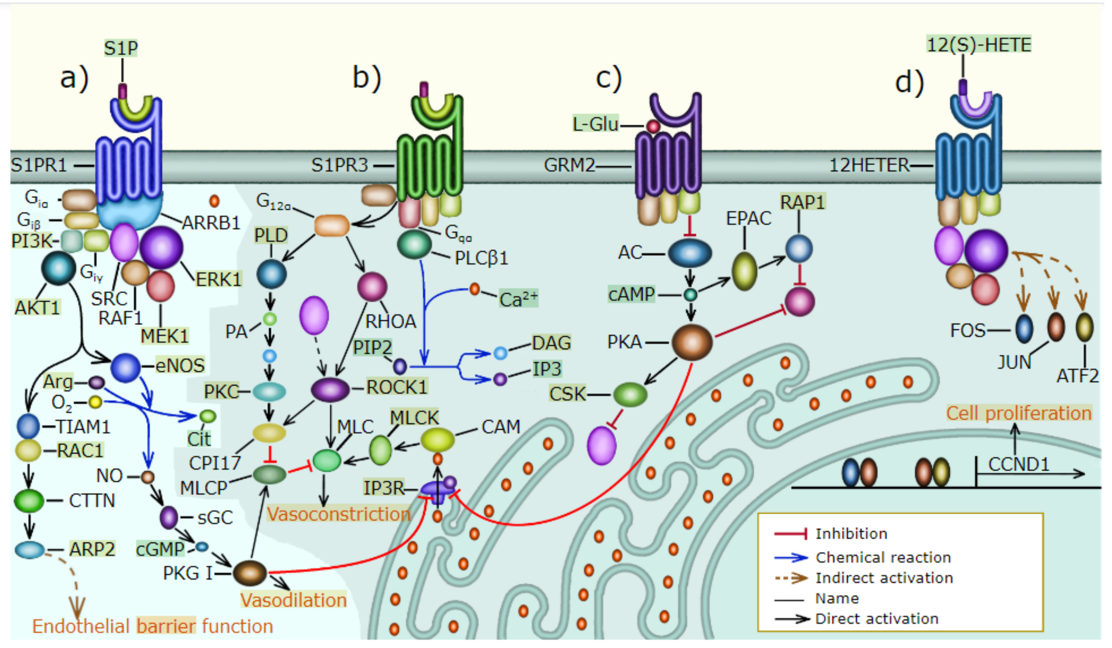


a) S1PR1 signaling, b) S1PR3 signaling, c) L-Glu signaling, d) 12(S)HETE signaling

### 3) Biological mechanisms associated with SRC

Semantic analysis of SRC

Biological Process — Table | Graph | Clustergram | Cloud

| GOID | GO Term | Associated genes | q-value |
|---|---|---|---|
| 70102 | **interleukin-6-mediated signaling pathway** | STAT3, IL6, SRC, CBL, STAT1 | 6.76e-7 *** |
| 42127 | **regulation of cell population proliferation** | STAT3, TNK2, SRC, JUP, STAT1 | 5.04e-3** |
| 3340 | **negative regulation of mesenchymal to epithelial transition involved in metanephros morphogenesis** | STAT1, CTNNB1 | 5.04e-3** |
| 2000635 | **negative regulation of primary miRNA processing** | IL6, STAT3 | 5.04e-3** |
| 48041 | **focal adhesion assembly** | PDPK1, CTTN, SRC | 6.40e-3** |

Biological process associated with SRC thorough statistical enrichment

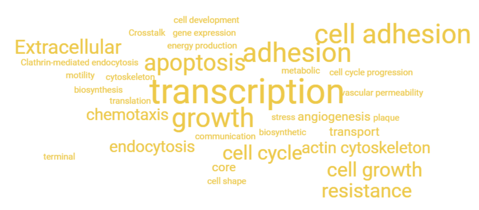


Phenotypes associated with SRC (Count per row word cloud)

### 4) How is SRC associated with IL6 signaling?

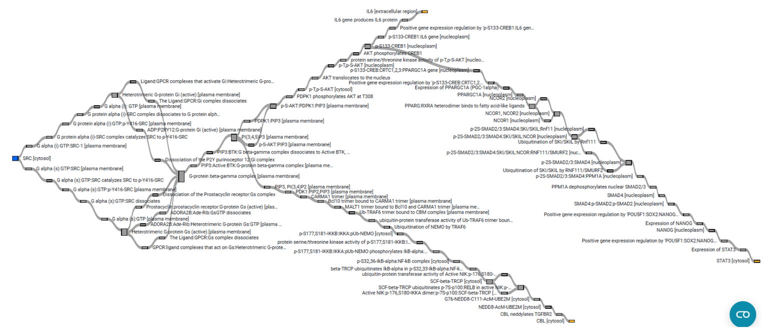

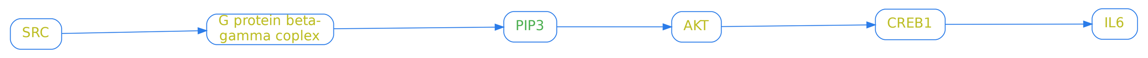


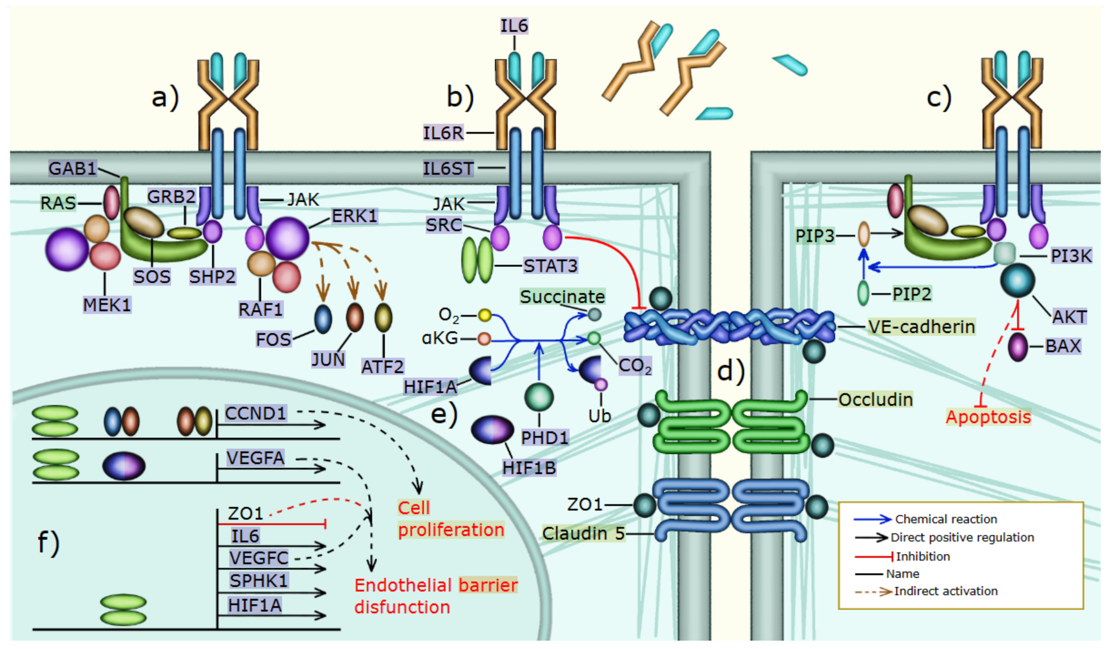



Singer, C. A., Lontay, B., Unruh, H., Halayko, A. J. & Gerthoffer, W. T. Src mediates cytokine-stimulated gene expression in airway myocytes through ERK MAPK. Cell Communication and Signaling 9, 1–5 (2011).

Simpson, C. E. et al. Cellular sources of interleukin-6 and associations with clinical phenotypes and outcomes in pulmonary arterial hypertension. European Respiratory Journal 55, (2020).

Steiner, M. K. et al. IL-6 Overexpression Induces Pulmonary Hypertension. Circ Res 104, 236 (2009).

Hirano, T., Ishihara, K. & Hibi, M. Roles of STAT3 in mediating the cell growth, differentiation and survival signals relayed through the IL-6 family of cytokine receptors. Oncogene 2000 19:21 19, 2548–2556 (2000).

Alsaffar, H., Martino, N., Garrett, J. P. & Adam, A. P. Interleukin-6 promotes a sustained loss of endothelial barrier function via janus kinase-mediated STAT3 phosphorylation and de novo protein synthesis. Am J Physiol Cell Physiol 314, C589–C602 (2020).

Kolliputi, N. & Waxman, A. B. IL-6 cytoprotection in hyperoxic acute lung injury occurs via PI3K/Akt-mediated Bax phosphorylation. Am J Physiol Lung Cell Mol Physiol 297, 6–16 (2009).


### 5) SRC and IL6 signaling is associated with Activin and BMP signaling


Tielemans, B., Delcroix, M., Belge, C. & Quarck, R. TGFβ and BMPRII signalling pathways in the pathogenesis of pulmonary arterial hypertension. Drug Discov Today 24, 703–716 (2019).


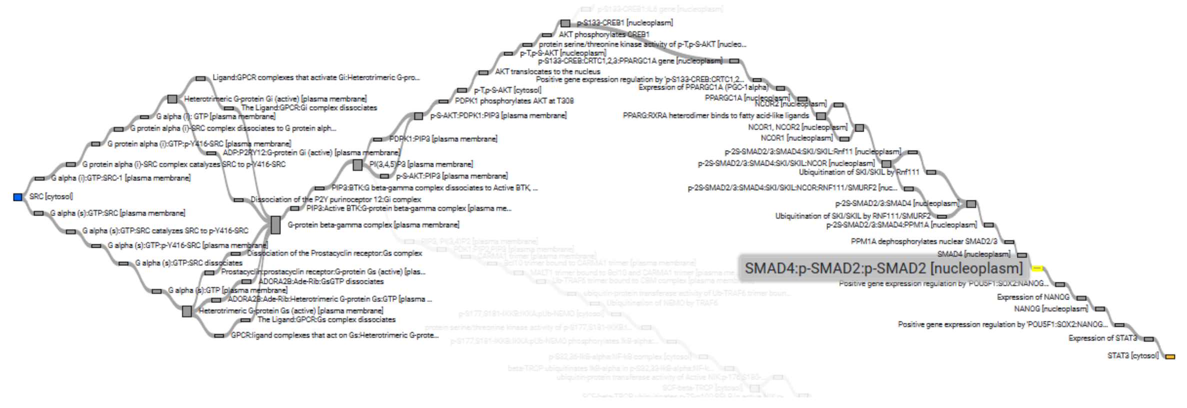


SMAD2 is present in the shortest path that connects SRC to STAT3

Twice expanded semantic analysis around SRCSRC x2

Biological Process — Table | Graph

Search: Activin

| GOID | GO Term | Associated genes |
|---|---|---|
| 32924 | **activin receptor signaling pathway** | SMAD2, SMAD3, TGFBR2 |
| 32925 | **regulation of activin receptor signaling pathway** | SMAD7 |

Biological Process — Table | Graph | Clustergram

Search: BMP

| GOID | GO Term | Associated genes |
|---|---|---|
| 30509 | **BMP signaling pathway** | SMAD4, SMAD2, RUNX2, LEF1, SMAD3, TGFB1, ENG, SMAD7, TGFBR3 |
| 30513 | **positive regulation of BMP signaling pathway** | SMAD4, SMAD2, FBXL15, ENG, NOTCH1 |
| 30514 | **negative regulation of BMP signaling pathway** | ABL1, SMAD4, PPARG, NOTCH1, SMAD7 |
| 71773 | **cellular response to BMP stimulus** | RUNX2, SOX9, SMAD4 |

### 6) Activin and BMP signaling in pulmonary hypertension

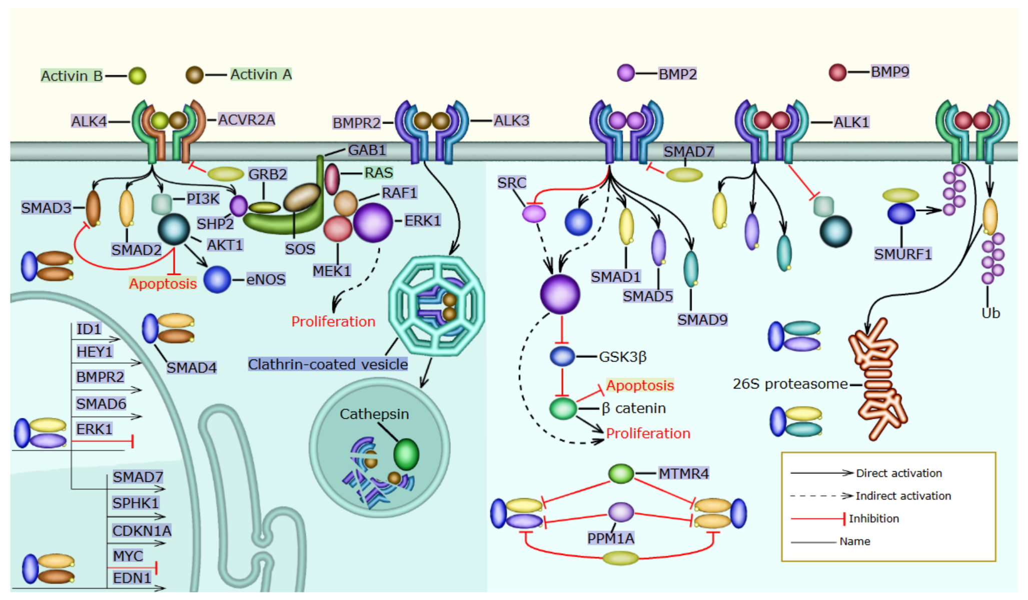



Yndestad, A. et al. Elevated levels of activin A in clinical and experimental pulmonary hypertension. J Appl Physiol 106, 1356–1364 (2009).

Ryanto, G. R. T. et al. An endothelial activin A-bone morphogenetic protein receptor type 2 link is overdriven in pulmonary hypertension. Nature Communications 2021 12:1 12, 1–14 (2021).

Humbert, M. et al. Sotatercept for the Treatment of Pulmonary Arterial Hypertension. New England Journal of Medicine 384, 1204–1215 (2021).



Created by Lifelike
lifelike.bio
