## Supplementary material for "A *Lifelike* guided journey through the pathophysiology of pulmonary hypertension - from measured metabolites to the mechanism of action of drugs": Pathways map

Why has Sotatercept only been used so far for PAH patients

### G protein signaling

Smooth muscle relaxation and protein kinase G
Smooth muscle relaxation occurs in a cAMP- and cGMP-dependent manner [28]. Stimulation of cAMP and protein kinase A (PKA) results from activation of adenylate cyclase, whereas cGMP and activation of protein kinase G (PKG) is mediated by nitric oxide (NO) binding to the heme group of a soluble guanylate cyclase or by a particulate (membrane-associated) receptor cyclase activated by atrial natriuretic factor [28].

Sph-1-P, derived from platelets and dephosphorylated on the cell surface, may induce the contraction of coronary artery smooth muscle cells through the S1P2/Rho signaling.

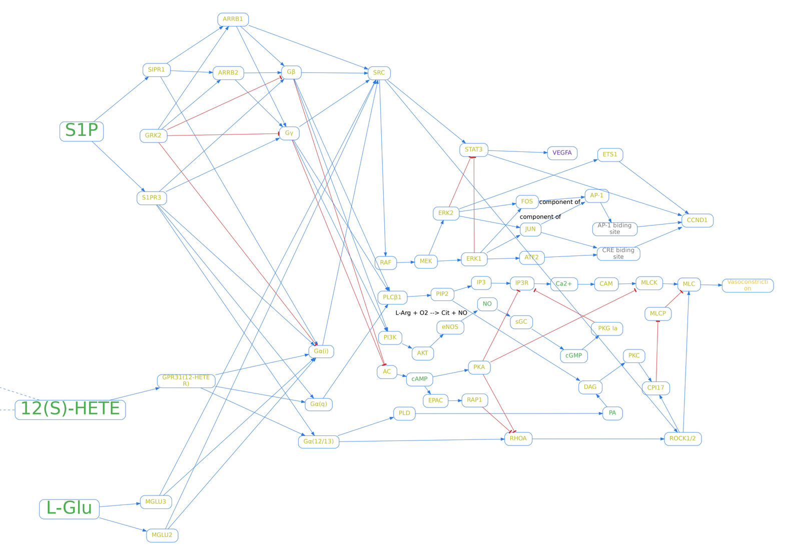


12(S)-HETE induces lymph endothelial cell retraction in vitro by upregulation of SOX18

Feed-back loops integrating RELA, SOX18 and FAK mediate the break-down of the lymph-endothelial barrier that is triggered by 12(S)-HETE

In endothelial cells, signaling trough mGlu1 or mGlu5 receptors may be protective during episodes of acute oxidative damage.

L-Glutamate has been shown to have a role in the maintenance of endothelial barrier (Langford et al., 1997).

Microvascular endothelial cells from human skin are positive for mGlu1, mGlu4, and mGlu5 (Collard et al., 2002).

An mGlu2/3 signal is present in the endothelial lining of the rat respiratory tissues (Gill et al., 2000).

#### Cell-Surface Receptors Transactivation Mediated by G Protein-Coupled Receptors

by Fabio Cattaneo [1,†], Germano Guerra [2,†], Melania Parisi [1], Marta De Marinis [1], Domenico Tafuri [3], Mariapia Cinelli [4] and Rosario Ammendola [1,*]

Tanimoto, T., Lungu, A. O., & Berk, B. C. (2004). Sphingosine 1-phosphate transactivates the platelet-derived growth factor β receptor and epidermal growth factor receptor in vascular smooth muscle cells. Circulation research, 94(8), 1050-1058.

Tanimoto T, Jin ZG, Berk BC. Transactivation of vascular endothelial growth factor (VEGF) receptor Flk-1/KDR is involved in sphingosine 1-phosphate-stimulated phosphorylation of Akt and endothelial nitric-oxide synthase (eNOS). J Biol Chem. 2002; 277: 42997–43001.

Endo A, Nagashima K, Kurose H, Mochizuki S, Matsuda M, Mochizuki N. Sphingosine 1-phosphate induces membrane ruffling and increases motility of human umbilical vein endothelial cells via vascular endothelial growth factor receptor and CrkII. J Biol Chem. 2002; 277: 23747–23754.

Nguyen, A. V., Wu, Y. Y., & Lin, E. Y. (2014). STAT3 and sphingosine-1-phosphate in inflammation-associated colorectal cancer. World journal of gastroenterology: WJG, 20(30), 10279.

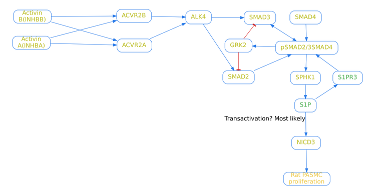


GPCR such as thrombin, angiotensin II, endothelin-1, and lysophosphatidic acid lead to the time-dependent generation of phosphorylated Smad2 (8., 9., 10., 11., 12.).

In inflammatory response whereby interleukin (IL)-6 production is abundant, STAT3-mediated pathways were found to promote the activation of sphingosine kinases (SphK1 and SphK2) leading to the production of S1P. Reciprocally, S1P encourages the activation of STAT3 through a positive autocrine-loop signaling

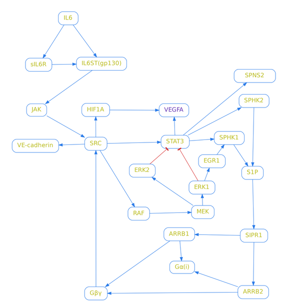


This article is highly recomended.:"S1P induction of keratinocyte migration as well as Smad3 phosphorylation required both S1P and TGFβ receptors (60). Furthermore, S1P activation of Smad3 was shown to be GPCR dependent. The notion that Smad3 activation was required for S1P induction of keratinocytes was further confirmed by the observation that neither S1P nor TGFβ could induce chemotaxis in keratinocytes from Smad3−/− mice. "

Fig. 2.
S1P induces the formation of an S1P receptor-TβRII receptor complex that forms a heteromer with TβRI resulting in phosphorylation of Smads 1, 2, and 3. P, phosphate; TGFβR, TGFβ receptor.

### IL6 signaling

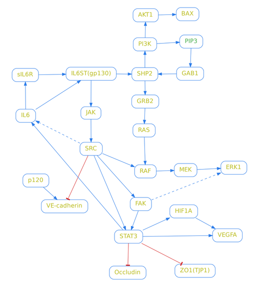


Fig. 9.
IL-6/JAK signaling in HUVEC promotes an early loss of barrier function that depends on MEK and Src activities (in blue) and a prolonged loss that depends on STAT3 phosphorylation at Y705 and de novo mRNA and protein synthesis (in red). Dashed lines represent translocation of the proteins to the nucleus or away from the cell-cell junctions. Gray lines represent pathways that are activated by IL-6 but are not required for IL-6-induced loss of barrier function.

FIGURE 1. INTERLEUKIN-6 CLASSICAL AND TRANS-SIGNALING. In the classical signaling, interleukin-6 (IL-6) binds to a membrane bound-IL-6 receptor (IL-6R) triggering the attachment and dimerization of glycoprotein 130 (gp130), followed by the activation of the non-receptor tyrosine kinase JAK that phosphorylates the STAT transcription factors. The activation of non-canonical signaling pathways, such as SHP-2/ERK, was also described. In the trans-signaling, soluble IL-6R (sIL-6R) was produced by shedding of the membrane bound IL-6R by ADAM10 or ADAM17. In human, but not in mice, sIL-6R was also produced by alternative splicing. IL-6 interacts with sIL-6R and the IL-6/sIL-6R complex binds a membrane bound gp130, activating the same signaling pathway described for classical signaling. Soluble gp130 (sgp130) was produced mainly by alternative splicing and also by shedding of the membrane bound gp130 by ADAM17 and

### Interaction between IL6 and TGFB signaling

JOURNAL ARTICLE

#### Smad3–STAT3 crosstalk in pathophysiological contexts

Yuka Itoh, Masao Saitoh, Keiji Miyazawa

*Acta Biochimica et Biophysica Sinica*, Volume 50, Issue 1, January 2018, Pages 82–90,
https://doi.org/10.1093/abbs/gmx118
**Published:** 13 November 2017 **Article history** ▾

Signal transduction through STAT3 and Smad2/3 STAT3 and Smad2/3 are activated via phosphorylation of a tyrosine or two serine residues, respectively, by ligand-stimulated receptor complexes, and inactivated by phosphatases (SHP2 or DUSP2 for STAT3 and PPM1A or MTMR4 for Smad3). Activated STAT3 and Smad2/3 translocate into the nucleus and regulate expression of target genes, including SOCS1/3 or Smad7. These proteins function as negative feedback regulators by

Mutual antagonism between Smad3 and STAT3 (A) STAT3 inhibits Smad2/3 signaling by inducing Smad7 expression. (B) STAT3 inhibits formation of the activated Smad complex by direct physical interaction with Smad3. (C) STAT3 recruits c-Ski and SnoN to the Smad3 promoter region and attenuates Smad3 expression. (D) Smad3 represses expression of the STAT3 gene, in cooperation with β2-SP, by inhibiting expression of ATF3/CREB2 and their recruitment to the STAT3 promoter region. (E) TGF-β inhibits STAT3 phosphorylation by repressing JAK2 expression and/or inducing SOCS1 expression.

Positive cooperativity of Smad3 and STAT3 (A) In the presence of Ras signaling, Smad3 disrupts the STAT3–PIAS3 complex by sequestering PIAS3 in response to TGF-β. Released STAT3 in turn induces Snail expression, possibly through non-canonical STAT3-binding element(s). (B) Activated STAT3 promotes nuclear localization of Smad3 and its recruitment to the Snail promoter to induce gene expression. It remains unclear whether STAT3 associates with the Snail promoter region. (C) Smad3 and STAT3 cooperatively induce CTGF expression in hepatic stellate cells. TGF-β induces the early-phase phosphorylation of STAT3 by TβRI-associated JAK1 (1), as well as the late-phase phosphorylation requiring Smad3, TβRI kinase activity, and de novo protein synthesis (2). Smad7 may be a mediator of the late-phase phosphorylation of STAT3. It remains to be determined how phosphorylation of STAT3 in the early-phase affects TGF-β signaling. In the late phase, Smad3 and STAT3 functionally cooperate to induce gene expression, possibly without a direct physical interaction.

### TGFB signaling

33 different ligand genes, including TGF-β1-3, after which the family is named, bone morphogenetic proteins (BMP), growth differentiation factors (GDF), activins, nodal, anti-Müllerian hormone (AMH), inhibins and leftys

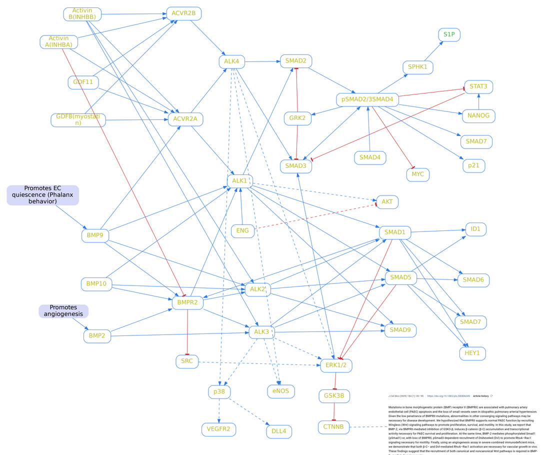


Once phosphorylated, these tyrosine residues serve as docking sites for numerous signaling molecules with either Src homology 2 (SH2) or phosphotyrosine binding (PTB) domains, such as Src and growth factor receptor binding protein 2 (Grb2).

In support of this view, Src was shown to phosphorylate TβRII on Tyr284, which serves as a docking site for the recruitment of growth factor receptor-bound protein 2 (Grb2) and Shc, thus linking TβRII to Ras and Erk activation (Galliher and Schiemann 2007).

PAH-Causing Mutations of BMPR-II on c-Src (Tyr418) Phosphorylation

hPASMC experienced a rapid decrease in c-Src phosphorylation at Tyr418 as compared with control. At as little as 10 min after stimulation, c-Src (Tyr418) phosphorylation was decreased by ~ 50% in response to BMP-2 and BMP-4, and by ~ 20% in response to BMP-7.

Overexpression of activin-A and -B in malignant mesothelioma – Attenuated Smad3 signaling responses and ERK activation promote cell migration and invasive growth

Created by Lifelike
lifelike.bio
