## Supplementary data for "A *Lifelike* guided journey through the pathophysiology of pulmonary hypertension - from measured metabolites to the mechanism of action of drugs"



November 21, 2023


### 1 Pathways in detail

#### 1.1 G protein-coupled receptor signaling

##### 1.1.1 In healthy people S1PR1 activity improves endothelial barrier function, promotes vasodilation, and enables normal PAEC proliferation.

In endothelial cells, S1PR1 is abundant, while S1PR2 and S1PR3 are present at a lower concentration [1, 2]. When S1P binds S1PR1, it leads to the formation of a protein complex that includes ARRB1,2, Gi/0$\alpha$, Gi$\beta$, and Gi$\gamma$ that causes the sequential recruitment, phosphorylation, and activation of SRC, RAF1, MEK1, MEK2, ERK1, and ERK2 [3, 4]. After that, ERK signaling promotes PAEC and PASMC proliferation by inducing Cyclin D1 expression. Cyclin D1 binds CDK4 or CDK6 to form a complex that is necessary for cell cycle progression through the G1 phase, and the G1/S transition. ERK1,2 is sufficient and necessary for the expression of Cyclin D1 [5]. ERK indirectly activates the transcription of the AP-1 components JUN and FOS, and the related protein ATF2. The Cyclin D1 promoter contains a consensus AP-1 site that is activated by FOS, JUN, and a cyclic AMP-response element (CRE) that can be activated by JUN and ATF2 [6]. Further, the Cyclin D1 promoter contains three Ets binding sites, ETS1 is activated by ERK1/2 and promotes bovine retinal endothelial cell proliferation [6, 7]. A normal plasma concentration of S1P (0.1 to 0.8 µM) prevents endothelial barrier permeability [8]. S1P-bound S1PR1 recruits Gi$\beta$ and Gi$\gamma$ that bind and activate PI3K. Then, PI3K generates PIP3 which recruits AKT1 to the cell membrane where it is activated. Subsequently, active AKT1 recruits the guanine exchange factor for RAC1(TIAM1). Later, AKT1 and TIAM1 bind SIPR1 forming a complex that activates RAC1. After that, RAC1 increases PI3K activity forming a positive feedback mechanism. Lastly, RAC1 inhibits the formation of RHOA-dependent stress fibers and recruits CORTACTIN to actin-nucleation complexes that contain ARP2 and ARP3, this causes the polymerization of actin filaments and EC spreading, therefore improving endothelial barrier function [2]. Additionally, AKT-1 activates eNOS by phosphorylating it on serine 1177. After that, eNOS catalyzes a reaction that converts L-arginine and $O_2$ into L-citrulline and NO, this increases endothelial NO production and secretion. Then, NO activates sGC, which converts GTP to cGMP. Later, cGMP activates PKG, which inhibits $Ca^{2+}$ import [9], promotes Ca2+ storage in the sarcoplasmic reticulum by inhibiting IP3R1 and activating SERCA [10]. Furthermore, PKG I induces MLCP function, which together with a decrease in cytosolic Ca2+ prevents MLC activation and VSMC contraction [11, 10].

##### 1.1.2 S1PR3 signaling promotes vasoconstriction and increases blood vessel permeability.

An elevated plasma concentration of S1P ( 5 µM) promotes vasoconstriction and vascular permeability. ECs and vascular SMCs express S1PR2, and S1PR3, however, both are far more prevalent in vascular SMCs. After S1P binds and activates S1PR2, or S1PR3, it signals through Gi/0, G12, G13, and Gq [8]. G12, and G13 signaling induces vasoconstriction by activating guanine nucleotide exchange factors (GEFs) that transform inactive GDP-RHOA into GTP-RHOA, which activates the Rho-kinases ROKI and ROKII. After that, ROK can directly phosphorylate MLC, and CPI-17, which inhibits MLCP and thus indirectly increases MLC activity [12, 13]. Furthermore, G12, and G13 can also activate the enzyme PLD that stimulates the hydrolysis of phosphatidylcholine resulting in phosphatidic acid (PA) production [14]. Then, PA is dephosphorylated and transformed into diacylglycerol (DAG), which promotes sustained protein kinase C (PKC) activity. PKC then phosphorylates and activates CPI-17 which increases MLC activity by inhibiting MLCP [15, 12]. S1PR3-activated Gi/0, Gq, G12, and G13 can stimulate phospholipase C (PLC) [16, 12, 8], however PLC activation is primarily mediated by Gq [17]. PLC$\beta$ with Ca2+ as a cofactor catalyzes a reaction that transforms phosphatidylinositol 4,5-bisphosphate (PIP2) into inositol 1,4,5-trisphosphate (IP3) and DAG [18]. As mentioned above, DAG promotes MLC activity. Additionally, when IP3 binds its receptor IP3R1 on the sarcoplasmic reticulum membrane (or the endoplasmic reticulum in ECs), it promotes the release of $Ca^{2+}$ into the cytoplasm that binds and activates Calmodulin (CAM). After that CAM activates MLCK [19], which phosphorylates and activates MLC, and that leads to VSMC contraction53.

##### 1.1.3 GRM2 and GRM3 signaling inhibits adenylate cyclase activity.

L-glutamate is involved in the endothelial response to oxidative damage and regulates endothelial barrier function [20]. The type II metabotropic L-Glu receptors GRM2 and GRM3 are expressed in rat pulmonary endothelial cells [21]. GRM2 and GRM3 are Gi/o-coupled receptors that inhibit Adenylate cyclase (AC) function leading to a lower concentration of cAMP, less PKA and C-terminal Src kinase (CSK) activity, causing an increase in SRC activity [22]. The decrease in cAMP hinders cAMP-inducible guanine-exchange factor for Rap (EPAC) function. EPAC enhances endothelial barrier function by increasing cortical actin and activating RAP1, which obstructs RHOA activation [23]. Additionally, PKA phosphorylates and inhibits MLCK in cultured bovine pulmonary arterial endothelial cells[24]. Further, PKA promotes vasodilation by inhibiting the IP3-dependent release of $Ca^{2+}$ from the sarcoplasmic reticulum [25]. Therefore, suppressing AC function promotes MLC-mediated cell contraction [23].

##### 1.1.4 12HETER signaling promotes cell proliferation.

12-(S)-HETE binds and activates the G protein-coupled receptor 12HETER (GPR31) that signals through Gi/0 to activate SRC, ERK1 and ERK2 [26]. Furthermore, 12(S)-HETE promotes PASMC proliferation through the activation of ERK [27]. Moreover, 12(S)-HETE causes EC retraction through the phosphorylation of cytoskeletal proteins in a path mediated by PKC [28]. Additionally, 12(S)-HETE leads to MLC2 phosphorylation to induce lymphatic endothelial barrier breaching, this process is mediated by G$\alpha$12/13 RHOA and ROCK [29].

### 1.2 Interleukin 6 Signaling

#### 1.2.1 IL6 signaling initiation.

In PH patients, SRC activity increases the expression of IL6 [30, 31]. The IL6 receptor IL6R is absent from the cell membrane of PAECs. However, in the plasma of PH patients, IL6 binds the soluble form of its receptor (sIL6R), and then both bind the coreceptor IL6ST (gp130), which is expressed by PAECs, forming a hetero-hexamer complex that initiates trans IL6 signaling. Under PAH conditions, PASMCs express both IL6, its receptor IL6R and the coreceptor IL6ST, therefore in PASMCs classical IL6 signaling is activated autocrinally [32, 33].

#### 1.2.2 ERK1, ERK2, and STAT3 are activated by IL6 and promote PAEC and PASMC proliferation.

IL6 promotes PAEC and PASMC proliferation by allowing cell cycle progression during the G1 phase and through the G1/S transition. The complex formed by IL6, IL6R, and IL6ST binds JAK, and either SHP2 [34] or SRC to activate RAF/MEK/ERK signaling (Figure 4 a) and STAT3 (Figure 4 b) [35, 36]. ERK1 and ERK2 increase the expression of the transcription factors FOS, JUN, and ATF2, which together with STAT3, bind to the promoter of CNND1 and increase the expression of Cyclin D1. Then, CDK4 or CDK6 and CyclinD1 form a complex that allows cell cycle progression through transcriptional regulation [6, 7] (Supplementary section 1.1.1.). Moreover, STAT3 activates the expression of MYC, PIM1, and PIM2 that also control the G1 to S transition [37]. Additionally, IL6 induces PASMC proliferation through M2 macrophage polarization. IL-6 promotes the differentiation of Th17 cells that secrete IL-17. CD4+ T cells including Th17 cells respond to IL-6 and IL-17 by secreting IL-21, which promotes M2 macrophage polarization. IL-21 and M2 macrophage markers were upregulated in the lungs of patients with idiopathic PAH. IL-21 receptor deficiency in mice leads to hypoxia-induced PH and prevents the accumulation of M2 macrophages in the lungs [38]. Lastly, the cytokines secreted by hypoxic M2 macrophages promote the proliferation of PASMCs [39].

#### 1.2.3 IL6-activated SRC and STAT3 increase endothelial barrier permeability.

After being activated by IL6 signaling, SRC and STAT3 increase endothelial permeability. SRC directly phosphorylates VE-cadherin, this phosphorylation is necessary for VEGFA and VEGFR2-induced endothelial permeability [40] and is involved in IL6-induced VE-cadherin internalization and loss of barrier function in PAECs [35]. Exposure to IL6 causes an increase in barrier permeability that lasts more than 24 hours. However, the effect of SRC, and VE-phosphorylation on endothelial barrier function is necessary only during the initial hours implying that IL6 increases endothelial permeability through other molecular mechanisms [41]. IL6 reduces the expression of VE-cadherin [41]. Further, IL6-induced STAT3 inhibits the expression of the tight junction components ZO1 and Occludin (Figure 3d, 3f) [42]. Furthermore, STAT3 activates the transcription of IL6, SPHK1, HIF1A, VEGFC, and VEGFA (Figure 3f).

#### 1.2.4 IL6 signaling and hypoxia activate the transcription of VEGFA.

Under hypoxic conditions, the ubiquitination and degradation of HIF1A is inhibited. Then, HIF1A forms a complex with HIF1B (Figure 3e) that binds the VEGFA promoter. When both STAT3 and HIF1A/HIF1B bind the VEGFA promoter, the expression of VEGFA is very high [43]. This constitutes an important connection between hypoxia, inflammation, and increased endothelial permeability that partially explains how IL6 contributes to the development of PH caused by hypoxia [44]. VEGFA has two effects that are important during PH progression, it promotes angiogenesis, and it increases endothelial barrier permeability through the phosphorylation of junction components [45], including ZO1, Occludin [46], and VE-cadherin. The downregulation of VE-cadherin is initiated when VEGFA binds neuropilin 1 (NRP1) [47] and VEGFR2 that activate SRC. Then SRC phosphorylates the guanine nucleotide-exchange factor VAV2, which activates the small GTPase RAC. After that, RAC activates p21-activated kinase (PAK) leading to the phosphorylation of a highly conserved motif within the intracellular region of VE-cadherin. This causes VE-cadherin to recruit ARRB2 promoting the internalization of VE-cadherin in clathrin-coated vesicles [48].

#### 1.2.5 IL6 inhibits apoptosis through AKT signaling.

In cultivated human vascular endothelial cells (HUVECs) AKT phosphorylation requires sIL6R, JAK, and PI3K [49]. The mechanism that allows IL6 signaling to activate PI3K is not fully known. However, it involves SHP2, IL6ST, and GAB1 [34]. IL6-activated AKT inhibits PAEC hyperoxia-induced apoptosis through the inhibition of the proapoptotic member of the Bcl-2 family BAX (Figure 3c) [50]. Additionally, AKT stimulates the expression of the inhibitor of apoptosis Survivin in endothelial cells [51].

#### 1.3 TGF signaling.

TGF$\beta$ signaling is initiated when a TGF$\beta$ family ligand dimer associates with two homodimers, one formed by type II -TGF$\beta$ receptor (T$\beta$RII) and the second comprising type I -TGF$\beta$ receptor (T$\beta$RI) [52, 53]. TGF$\beta$ ligand binding causes T$\beta$RIIs to phosphorylate four threonine or serine residues in the GS domain (TTSGSGSG motif) of T$\beta$RIs resulting in the activation of T$\beta$RIs [54]. The activated T$\beta$RIs can recruit, phosphorylate, and activate receptor-regulated SMADS (R-SMADs) [54]. After that, Common partner SMADs (SMAD4) bind two phosphorylated R-SMADs forming a complex that is transported from the plasma membrane into the nucleus where it regulates the transcription of several target genes. After TGF$\beta$ ligands activate T$\beta$RIIs and T$\beta$RIs, the latter phosphorylate and activate SMADs. Additionally, active T$\beta$RIIs and T$\beta$RIs modulate p38, JNK, AKT, ERK, and RHO signaling [55]. The mechanism that allows TGF$\beta$ ligands to activate ERK signaling requires the following steps: Ligand binding causes the phosphorylation of T$\beta$Rs that associate with a docking protein like SHP2, SRC, or SHCA. The docking protein then recruits a complex formed by GRB2 and SOS from the cytoplasm to the plasma membrane. Subsequently, SOS activates RAS by catalyzing the exchange of GDP for GTP. After that, GTP-bound RAS binds RAF leading to the sequential activation of RAF, MEK and ERK [55] (Figure 5a). After a ligand from the BMP or Activin branch of the TGF$\beta$ family associates with them, T$\beta$RIIs and T$\beta$RIs can bind PI3K, which phosphorylates and activates AKT [55] (Figure 5a). AKT activity has several functions which are relevant for PH, it activates eNOS that increases NO production [56]. Also, AKT inhibits BAD and BAX-mediated apoptosis [51]. Under certain conditions, TGF$\beta$ signaling can also inhibit AKT signaling. One possible molecular mechanism for this inhibition involves the docking protein SHCA, which reduces the expression of AKT and eNOS [57]. Additionally, in hematopoietic cells, TGF$\beta$ can reduce AKT signaling activity through the Smad-dependent expression of the lipid phosphatase SHIP [58]. TGF$\beta$ signaling is modulated through a diverse group of molecular mechanisms [59]. LEFTYA and LEFTYB function as competitive inhibitors for TGF$\beta$ ligands [60]. Additionally, certain ligands that have a higher binding affinity for T$\beta$RIIs prevent other ligands from associating with them [61]. BAMBI prevents the formation of functional receptor complexes by associating with T$\beta$RIs and T$\beta$RIIs. Furthermore, BAMBI collaborates with SMAD7 to prevent R-SMAD activation [62]. SMAD6, and SMAD7 are inhibitory SMADs (I-SMADs) that bind T$\beta$RIs preventing R-SMAD activation. Additionally, I-SMADs recruit ubiquitin E3 ligases, like Smad ubiquitin regulatory factors (Smurfs), which mediate the ubiquitination and degradation of active R-SMADS, T$\beta$RIs, and T$\beta$RIIs. Moreover, Smurfs and Salt-inducible kinase (SIK) enhance the interaction between I-SMADs and T$\beta$RIs to reduce R-SMAD activation. Furthermore, I-SMADs associate with active R-SMADs and prevent R-SMADs from associating with SMAD4. Also, I-SMADS present in the nucleus inhibit the transcription of R-SMAD-regulated genes [59]. What's more, the phosphatase PPM1A deactivates R-SMADs and promotes proteosome-mediated degradation of R-SMADs [63]. Lastly, the phosphatase MTMR4 dephosphorylates and deactivates R-SMADS within early endosomes [64].

##### 1.3.1 Activin signaling is overactivated in PH patients, promotes excessive vascular cell proliferation, and downregulates BMP signaling.

The TGF$\beta$ family ligands activin A, and activin B initiate activin signaling. The concentration of activin A is more elevated than normal in PH patients and is significantly correlated to mortality 99. Additionally, pulmonary microvascular endothelial cells (PMECs) isolated from idiopathic PAH patients exhibit augmented activin A expression [65]. Moreover, the PAECs of the same group of patients overexpressed activin B [65]. Further, the concentration of phosphorylated SMAD2 and SMAD3, and the immunoreactivity of ACVR2B, ALK2, ALK4, and ALK5 was more elevated than normal in PH patients [66]. Activin A and activin B associate with high affinity to the T$\beta$RIIs ACVR2A, ACVR2B, and BMPR2 [61], which are expressed in human pulmonary microvascular ECs [67].

**Normally SMAD-dependent activin signaling inhibits cell proliferation, increases the production of S1P, and modulates Activin and BMP signaling.** The T$\beta$RIs ALK4, ALK2, ALK3 and ALK6 compete to bind ACVR2A on the cell membrane [68]. Moreover, activin A and ALK4 are expressed in pulmonary endothelial cells [69] and the complex formed by ACVR2A and ALK4 transduces activin A signaling to activate SMAD2 and SMAD3 [68, 70]. SMAD2 and SMAD3 bind SMAD4 forming complexes activate the transcription of CDKN1A (p21) and inhibit the expression of MYC to prevent the proliferation of cultured human umbilical vein endothelial cells (HUVECs) [71, 72]. Additionally, SMAD2 and SMAD3 increase the production of S1P by activating the expression of SPHK1 in rat PASMCs, the increase in S1P production is normally beneficial [73]. Also, SMAD2 and SMAD3 modulate Activin and BMP signaling by stimulating the transcription of SMAD7 [74]. Further, the complex formed by SMAD2, SMAD3 and SMAD4 can activate the transcription of SMAD3 forming a positive feedback mechanism that amplifies canonical activin signaling [75].

**SMAD-independent activin signaling promotes EC proliferation, inhibits apoptosis, and downregulates BMP signaling.** Activin also signals through SMAD-independent pathways. Activin A induces ERK signaling in fibroblasts

[76]. Also, activin B activates ERK signaling in adipose [77] and bone marrow-derived [78] mesenchymal stem cells, which are closely related to ECs through the endothelial-to-mesenchymal transition. Further, ERK signaling stimulates PAEC and PASMC proliferation (Supplementary section 1.2.2.). Activin A also causes the sequential activation of PI3K, AKT and eNOS in endothelial cells from the atrioventricular canal during early development [56]. Additionally, AKT signaling inhibits PAEC apoptosis (Supplementary section 1.2.5.). Notably, AKT directly associates with SMAD3 preventing its T$\beta$RI-mediated phosphorylation and nuclear translocation [79, 55]. Activin signaling promotes the internalization and degradation of BMPR2. When activin A binds BMPR2 it leads to clathrin-mediated endocytosis of BMPR2. Then the vesicles containing BMPR2 fuse with lysosomes that contain Cathepsins and degrade BMPR2 resulting in a lower concentration of BMPR2 in the membrane of in PAECs [65].

#### 1.3.2 The effect of BMP signaling on PH.

The BMP branch of the TGF$\beta$ family of ligands includes BMP2, BMP4, BMP6, BMP9, and BMP10. Normally, BMP9 and BMP10 are present in the blood serum at a sufficient concentration to elicit BMP signaling ( 0.5–15 ng/mL). BMP2, BMP4, and BMP6 are present at a low concentration and their expression is carefully regulated spatially and temporally leading to the formation of dynamic gradients of these ligands. Notably, some endothelial cells express BMP2, BMP4, and BMP6 [80]. Extracellular antagonists including Chordin, Noggin, Gremlin, and MGP modulate BMP signaling. Noggin and Gremlin bind and inhibit BMP2, BMP4, and BMP7, while Chordin binds and inhibits BMP4 [80].

**BMP2 signaling is inhibited in PH patients.** Normally, BMP2 signaling prevents excessive pulmonary vascular cell proliferation, preserves endothelial barrier function, and promotes vasodilation by deactivating SRC and stimulating the expression of eNOS. In a mouse model of hypoxia-induced PH, the expression of BMP2 and BMP4 is upregulated in the peripheral pulmonary vasculature after exposure to hypoxia. Additionally, heterozygous null Bmp2+/- mice develop more severe PH than wild-type mice under hypoxic conditions [81]. BMP2 mostly associates with the T$\beta$RII BMPR2, which is normally expressed in PAECs and to a lesser degree in PASMCs [82]. Heterozygous Bmpr2 mutations are common in familial PAH patients. In mice, EC-specific Bmpr2 deletion spontaneously causes PH, and increases the severity of hypoxia-induced PH [81, 82]. The T$\beta$RI with the highest BMP2 binding affinity is ALK3 (BMPR1A) [83] and the formation and activation of the complex composed of BMP2, BMPR2 and ALK3 leads to the phosphorylation and activation of SMAD1, SMAD5, and SMAD9 [80]. The expression of BMPR2 and ALK3 is markedly reduced in the pulmonary vasculature of PH patients [84]. BMP2 has several beneficial effects that reduce the severity of PH. During the progression of hypoxia-induced PH, the activity of SMAD1, SMAD5, and SMAD9 increases. Then, the BMP-activated SMADs induce the transcription of the inhibitors of differentiation ID1, and ID3, that limit excessive PAEC and PASMC proliferation [85]. Additionally, SMAD1, SMAD5, and SMAD9 stimulate the transcription of SMAD6 and SMAD7, this is one of the molecular mechanisms that allows precise special and temporal modulation of TGF$\beta$ signaling [86, 87]. Moreover, BMP-activated SMADs inhibit the expression of ERK1 and ERK2, this is an important step during the endothelial-to-hematopoietic transition and inhibits vascular cell proliferation. Inhibition of SMAD1 and SMAD5 or overexpression of ERK1 caused artery enlargement and reduced the number of hematopoietic stem cells that form in 24 hpf Zebrafish embryos [88]. Furthermore, SMAD1 and SMAD5 induce the transcription of HEY1 [87], which is an important component of the molecular mechanism that regulates the endothelial-to-mesenchymal transition [89] and pulmonary fibrosis [90], two EndMT-related and important processes for PH [91, 92]. BMP2 activates ERK phosphorylation. Then, ERK1 deactivates GSK3$\beta$ preventing the inhibition of $\beta$-catenin, which promotes PAEC proliferation and inhibits excessive PAEC apoptosis [93]. Additionally, BMP2 signaling preserves endothelial barrier function and promotes vasodilation by activating eNOS [81, 94]. Moreover, BMP2 signaling prevents excessive pulmonary vascular cell proliferation by downregulating SRC activity. BMP2 activates BMPR2 and that decreases SRC phosphorylation at Tyrosine 418 in PASMCs [95].

**BMP4 inhibits PASMC apoptosis.** In contrast to BMP2 and BMP9, the effects of BMP4 signaling are mostly detrimental to PH patients. The expression of BMP4 is upregulated in the peripheral pulmonary vasculature of mice after exposure to hypoxia. Additionally, heterozygous null Bmp4+/- mice develop less severe PH than wild-type mice under hypoxic conditions [81]. BMP4 associates with the T$\beta$RII BMPR2 and the T$\beta$RI ALK3 leading to the phosphorylation and activation of SMAD1, SMAD5, SMAD9 and PI3K [81, 96]. In human PASMCs BMP4 signaling inhibits the cleavage of procaspase-3 and the expression of caspase-3, these BMP4 effects require AKT activity. Additionally, ALK3 deletion in mice PASMCs prevented the muscularization of distal pulmonary vessels caused by hypoxia [84]. Therefore, BMP4 signaling contributes to the excessive PASMC cell proliferation observed in PH patients [96]. Further, BMP4 causes vascular calcification [97, 98], and promotes the differentiation of endothelial cells into osteoblasts [89].

**BMP9 promotes EC quiescence, inhibits apoptosis, prevents excessive PAEC proliferation, preserves endothelial barrier function, and has a beneficial effect on animal models of PH.** BMP9 inhibits EC migration and proliferation

| Entry | Edge type |
|---|---|
| 1 | activeUnitOf |
| 2 | candidateOf |
| 3 | catalystOf |
| 4 | catalyzes |
| 5 | componentOf |
| 6 | input |
| 7 | memberOf |
| 8 | output |
| 9 | regulates |
| 10 | regulatorOf |
| 11 | repeatedUnitOf |
| 12 | requiredInput |

Table 1: Edge types included in algorithmic analyses performed on the Reactome graph database.

and usually forms a signaling complex with the T$\beta$RII BMPR-II and the T$\beta$RI ALK1, which is expressed mainly in ECs. This complex causes the phosphorylation and activation of SMAD1, SMAD5 and SMAD9 [99]. In human PAEC cultures, BMP9 dose-dependently increases the expression of BMPR2 and ID1, and upregulates SMAD1, SMAD5, and SMAD9 phosphorylation [100]. Additionally, in human PAECs BMP9 prevents JNK phosphorylation, caspase-3 cleavage, and apoptosis through BMPR2. Moreover, in rats, BMP9 reversed PH induced by monocrotaline or by a combination of hypoxia and Sugen. Notably, BMP9 prevented the muscularization of small vessels in the lung and the enlargement of the right heart ventricle caused by hypoxia and Sugen [100]. Also, in human PAECs, BMP9 can associate with ACVR2A and ALK1 resulting in the phosphorylation of SMAD2 but not SMAD3. The activity of SMAD2 is necessary for the expression of E-selectin and IL8 caused by BMP9 [101]. Further, heterozygous loss of function mutations in the gene GDF2 that encodes BMP9 are likely the cause of PH in some patients [102]. Loss of BMPR2 function in human PAECs reverses the beneficial effects of BMP9, in this context BMP9 promotes human PAEC proliferation and increases angiogenesis in mice [103].

### 2 Supplementary Tables and Figures

For more information on Supplementary Material and for details on the different file types accepted, please see the Supplementary Material section of the Author Guidelines.

Figures, tables, and images will be published under a Creative Commons CC-BY licence and permission must be obtained for use of copyrighted material from other sources (including re-published/adapted/modified/partial figures and images from the internet). It is the responsibility of the authors to acquire the licenses, to follow any citation instructions requested by third-party rights holders, and cover any supplementary charges.

#### 2.1 Tables

### 2.2 Figures

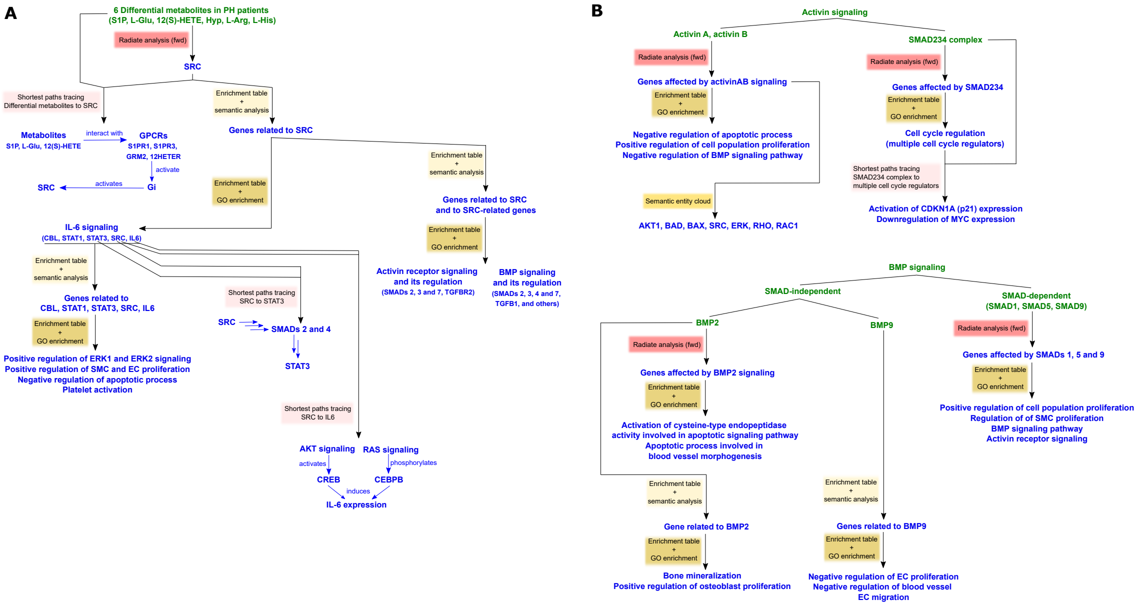


Figure 1: Workflows illustrating the combinatorial application of *Lifelike*'s data analytics methods to identify key biological processess in pulmonary hypertension. The analysis workflow was initiated either with six differential metabolites detected in PH patient plasma (A) and led us to analyze Activin and BMP signaling (B).
